# Predicting subnational incidence of COVID-19 cases and deaths in EU countries

**DOI:** 10.1101/2023.08.11.23293400

**Authors:** Alexis Robert, Lloyd AC Chapman, Rok Grah, Rene Niehus, Frank Sandmann, Bastian Prasse, Sebastian Funk, Adam J Kucharski

**Affiliations:** Centre for Mathematical Modelling of Infectious Diseases, London School of Hygiene and Tropical Medicine, London, UK; Department of Mathematics and Statistics, Lancaster University, Lancaster, UK; European Centre for Disease Prevention and Control (ECDC), Stockholm, Sweden

**Author notes:** These authors are joint first authors (Alexis Robert, Lloyd AC Chapman).

## Abstract

**Background:** Recurring COVID-19 waves highlight the need for tools able to quantify transmission risk, and identify geographical areas at risk of outbreaks. Local outbreak risk depends on complex immunity patterns resulting from previous infections, vaccination, waning and immune escape, alongside other factors (population density, social contact patterns). Immunity patterns are spatially and demographically heterogeneous, and are challenging to capture in country-level forecast models.

**Methods:** We used a spatiotemporal regression model to forecast subnational case and death counts and applied it to three EU countries as test cases: France, Czechia, and Italy. Cases in local regions arise from importations or local transmission. Our model produces age-stratified forecasts given age-stratified data, and links reported case counts to routinely collected covariates (test number, vaccine coverage..). We assessed the predictive performance of our model up to four weeks ahead using proper scoring rules and compared it to the European COVID-19 Forecast Hub ensemble model. Using simulations, we evaluated the impact of variations in transmission on the forecasts. We developed an open-source RShiny App to visualise the forecasts and scenarios.

**Results:** At a national level, the median relative difference between our median weekly case forecasts and the data up to four weeks ahead was 25% (IQR: 12-50%) over the prediction period. The accuracy decreased as the forecast horizon increased (on average 24% increase in the median ranked probability score per added week), while the accuracy of death forecasts remained stable. Beyond two weeks, the model generated a narrow range of likely transmission dynamics. The median national case forecasts showed similar accuracy to forecasts from the European COVID-19 Forecast Hub ensemble model, but the prediction interval was narrower in our model. Generating forecasts under alternative transmission scenarios was therefore key to capturing the range of possible short-term transmission dynamics.

**Discussion:** Our model captures changes in local COVID-19 outbreak dynamics, and enables quantification of short-term transmission risk at a subnational level. The outputs of the model improve our ability to identify areas where outbreaks are most likely, and are available to a wide range of public health professionals through the Shiny App we developed.

## Introduction

Local dynamics of COVID-19 depend on a combination of the level of immunity in a population, which in turn depends on previous incidence, vaccine uptake, and the immune escape properties of the currently circulating SARS-CoV-2 variants, and other factors such as population density, social contact patterns, migration patterns and adherence to public health measures. Most countries in Europe have been affected by repeated COVID-19 waves since March 2020, and have implemented extensive vaccination campaigns in order to reduce the health impact of these waves. This led to high national levels of both natural and vaccine-induced immunity (1). Before the emergence of the Delta and Omicron variants of concern (VOCs), this immunity provided considerable and durable protection against severe outcomes (hospitalisation and death) and some transient protection against infection (2). The emergence of new VOCs, and the extent of their immune escape, along with the waning of efficacy observed for a two-dose vaccine course, precipitated the expansion and acceleration of “booster” vaccination campaigns, further shifting the immune landscape of the population. Overall immunity to the Omicron BA.1 variant rose quickly after its emergence due to the unprecedentedly large wave of BA.1 cases that occurred throughout Europe in late 2021-early 2022, and vaccine booster campaigns. Since then, repeated emergence of Omicron subvariants with high levels of immune escape has led to multiple waves of infections, with lower case burden than the first BA.1 wave.

Currently, the overall level of immunity against infection is high compared to the early phase of the COVID-19 pandemic. The immune landscape of the population is spatially heterogeneous due to considerable variation in infection histories (e.g. multiple reinfections with different variants) and differences in vaccine uptake and timing across the population. If overall immunity rises, this immune landscape should result in the risk of outbreaks becoming more spatially and demographically heterogeneous, with the potential for distinct identifiable outbreaks. Therefore, there is interest from public health agencies in forecasting short-term incidence at a subnational level, and in different age groups, to anticipate large localised spikes in case numbers and local pressure on health care systems. Predicting such spikes in health care demand will become increasingly relevant as time moves forward if COVID-19 becomes more seasonal and influenza-like in its dynamics. This project was developed in collaboration with the European Centre for Disease Prevention and Control (ECDC), with the objective of developing a statistical framework for forecasting local case and death incidence in a range of European countries, and visualisation tools to communicate the predictions. The forecasts and scenario analysis can then be used to optimise planning and allocation of resources.

There have been numerous attempts to model subnational incidence of COVID-19 over the course of the pandemic. These have tended to fall into two broad categories: mechanistic “spatial” susceptible-exposed-infectious-recovered transmission models and statistical time series/spatiotemporal models. Most time series and transmission models have treated subnational regions as independent (fitted the model and made predictions separately for each region) without accounting for spatiotemporal correlations in incidence between regions (3–7), despite these being strong (8–10). A limited number of transmission models have instead treated subnational regions as connected sub-populations via a metapopulation approach, and used geographical distance or mobility/commuting data to parametrise connectivity between regions (11–16). These models become increasingly complex and hence slow to generate predictions as the number of affected regions increases, and have therefore received limited use for real-time forecasting during the pandemic. Time series models and spatiotemporal statistical models, on the other hand, have been used extensively and successfully to forecast future incidence at national and subnational levels (6,7,17). In the latter category, we focus on Endemic-Epidemic models, a flexible class of spatiotemporal statistical models that can be used to link changes in incidence to recent case numbers and the effect of various different covariates, on which the framework used in this paper is based. Endemic-Epidemic models have already been employed during the pandemic to understand and forecast spatiotemporal spread of COVID-19 at a subnational level (18–21) and at a national level in Africa (22), and to assess the impact of non-pharmaceutical interventions (NPIs), including lockdowns and border closures (23–26) (see (27) for a review). Their superiority to time series models that assume independence between regions has been demonstrated on data from northern Italy (28). However, they have not, as yet, been applied to forecast subnational incidence across multiple countries.

In this paper, we present a flexible modelling framework to capture subnational, age-stratified case dynamics of COVID-19, alongside a publicly available RShiny App to visualise the results and forecasts generated. The framework is used to predict subnational incidence of COVID-19 cases and deaths from routinely collected, publicly available surveillance data, and forecast the impact of changes in transmission on short-term dynamics (e.g. due to changes in behaviour or transmissibility). The framework can be run with an age-stratified model if local, age-stratified data is available. Public health professionals can use the RShiny App to visualise the forecasts of reported cases and deaths, and the predicted incidence under different scenarios, to get a full picture of the anticipated short-term burden of COVID-19 in local areas. We apply the framework to forecast cases and deaths at a NUTS-3 (Nomenclature of Territorial Units for Statistics 3) level in France, Czechia and Italy as test cases, and evaluate its ability to predict subnational incidence up to four weeks ahead.

## Methods

### Spatiotemporal modelling framework for reported cases

We model subnational COVID-19 case counts using the Endemic-Epidemic spatiotemporal modelling framework (29), as implemented in the *surveillance* R package (30). Endemic-Epidemic models provide a flexible framework for relating current incidence of cases to recent case incidence and to imported cases, accounting for the influence of other factors (i.e. covariates) on these relationships. They decompose local incidence in region *i* at time *t* into three components:

- An autoregressive component that quantifies the number of new infections expected from cases reported in region *i* at previous time steps.
- A neighbourhood component that quantifies the number of new infections expected from cases reported in regions around region *i* in previous time steps, which depends on the connectivity (or the human mobility) between regions.
- An endemic component that quantifies the background number of new infections occurring in region *i* at time *t*, independent of the current level of transmission, representing importations from regions outside those included in the study (or cases that could not be linked to the mechanistic components).

Endemic-Epidemic models are able to integrate multiple data sources from disease surveillance activity to link case incidence and various covariates. Each component is independently impacted by each covariate. Since we introduce various covariates and controls in our model, we merge the autoregressive and neighbourhood component into an epidemic component to reduce the number of parameters estimated, and avoid identifiability and convergence issues.

Early versions of the Endemic-Epidemic models only included dependence of current cases on cases in the previous time step, but they have since been extended to account for distributed lags via the *hhh4addon* R package (31–33). This allows for a more faithful representation of the impact of recent incidence on current case number, for instance by aligning the lag distribution to the serial interval of the pathogen.

The link between indicators of immunity and risk of SARS-CoV-2 infection is complex and unstable, due to waning of vaccine and infection-induced immunity, and emergence of variants able to escape immunity. The flexibility of the Endemic-Epidemic framework therefore makes it well suited to capture local dynamics of COVID-19 cases, while fully mechanistic frameworks may be too complex to parametrise. Depending on data availability, the model can be stratified by age.

### Non-age-stratified model

The total number of cases (over all age groups) in region *i* at time *t, Y*_*it*_, conditional on the number of cases in the same region and neighbouring regions in the previous *p* time steps, is modelled as negative binomial:

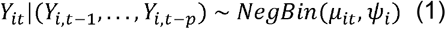

with conditional mean *μ*_*it*_ and dispersion parameter *ψ*_*i*_ such that 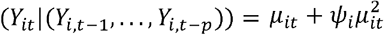

The mean is given by

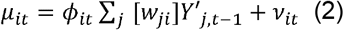

where 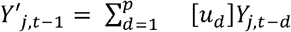 is the transmission potential from recent cases in region *j*, with [*u*_*d*_] the normalised lag weight for cases *d* days ago. We set *p*= 20 days and use a custom composite serial interval distribution for *u*_*d*_ that accounts for missing infection generations (34), with a mean and standard deviation for the first infection generation of 5 days and 1.5 days respectively, based on the estimated serial interval for SARS-CoV-2 for pre-Omicron variants (35–37), and 80% of the composite serial interval assumed to reflect direct transmission (without missing infections) (Supplementary Figure 1). The predictors for the epidemic (combined autoregressive and neighbourhood) and endemic components, *ϕ*_*it*_ and *v*_*it*_, determine the number of cases stemming from each component and are assumed to depend on log-linear component-specific predictors:

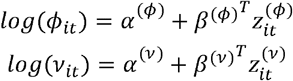

The predictors are independently impacted by different covariates, 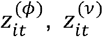, i.e. a covariate may be associated with fewer imported cases (endemic component *v*_*it*_), but have little impact on the spread of the virus in and between the regions (epidemic component *ϕ*_*it*_). The association between the covariates and the local number of cases expected is quantified by regression coefficients, *β*^(*ϕ*)^, *β*^(*v*)^, estimated through a maximum-likelihood approach. The weights *w*_*ji*_ in the neighbourhood component quantify the degree of connectivity between region *i* and surrounding regions *j*. We use a power-law model for the neighbourhood weights:

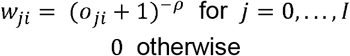

where *o*_*ji*_ is the adjacency order between regions *i* and *j* and *ρ* is a decay parameter to be estimated. The adjacency order is defined as the minimum number of borders that must be crossed to get from *i* to *j*: the adjacency degree is equal to 1 between neighbours, 2 between neighbours of neighbours, and so on. We normalise the weights, such that 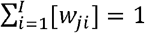, where, *I* is the maximum adjacency order considered, which we take to be 5.

### Age-stratified model

For countries in which age-stratified subnational case count data is available, we implement an age-stratified version of the model above using modified code from the *hhh4contacts* R package (38,39). For this model, the number of cases in age group in region *i* at time *t, Y*_*ait*_, conditional on the numbers of cases in the previous *p* time steps in the same and other age groups in region *i* and the surrounding regions is modelled as negative binomial:

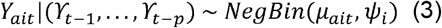

with conditional mean

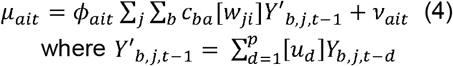

and overdispersion parameter *ψ*_*i*_ >0 such that 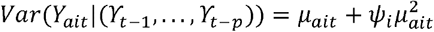, where 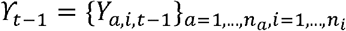 with *n*_*a*_ age groups and *n*_*i*_ regions, and *c*_*ba*_ is the mean number of daily contacts in age group *a* of an individual in age group *b*. We stratify the population into *n*_*a*_ =9 age groups (0-9, 10-19,…, 70-79, 80+ years). We use age-structured contact matrices from country-specific pre-pandemic contact surveys for *c*_*ba*_ where available, e.g. for France (40), and synthetic contact matrices estimated from contact survey and demographic data for countries in which no nationally representative contact surveys have been conducted, e.g. for Czechia (41). We used age-stratified intercepts in the epidemic component.

### Covariates

We incorporate various covariates in 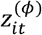 and 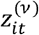 in the log-linear predictors for *ϕ*_*it*_ and *v*_*it*_. Thefull model equations are given in the Supplementary Material (see Model equation for each country and Supplementary Table 1). The covariates were picked based on potentially having had an effect on transmission and importation risk, or having an effect in future. We compare the forecasts obtained with the full model with covariates against a baseline model without covariates in the Supplement.

We included the same set of covariates in the endemic component in the age-stratified and non-age-stratified models:

- population: the total population of region *i*
- urban/rural status: binary covariates for whether the NUTS-3 region *i* is classified as urban, intermediate urban, intermediate rural or rural.
- seasonality: sinusoidal terms with annual periodicity to account for seasonal effects on importations. The amplitude and offset of the seasonality function are estimated by the model.
- number of cases in the WHO European region over the last month.

These covariates cover the impact of demographic characteristics and transmission in neighbouring countries on the background number of cases in each region (and age group in the age-stratified version of the model). The covariate specifications of the epidemic component depend on the availability of age-stratified data as follows:

- population:
  - age-stratified: two covariates:
    - total population of region *i*
    - proportion of the population of age group *a* in region *i*.
  - non age-stratified: one covariate: the total population of region *i*.
- testing:
  - age-stratified: two covariates:
    - proportion of the population in the country tested in the last 2 weeks.
    - if local age stratified testing data is available: proportion of population of age group *a* in region *i* (if local testing data is available, national otherwise) tested in the last two weeks. Otherwise, proportion of population of age group *a* in the country tested in the last two weeks.
  - non age-stratified: one covariate: the proportion of the population in the country tested in the last 2 weeks.
- vaccination coverage: the proportion of the population in region *i* who received their second dose in the last 120 days, or who have received three or more doses (for each age group in the age-stratified model)
- cumulative incidence: the cumulative incidence of cases between the start of the fitting period and a month ago in region *i* (for each age group in the age-stratified model)
- recent incidence: the cumulative incidence of cases in the past month in region *i* (for each age group in the age-stratified model)
- variant: two binary indicator variables for whether the proportion of sequenced cases that were Delta or an Omicron variant was higher than 30% (in all three countries, the Delta covariate is equal to one in late 2021, and the Omicron covariate is equal to one from January 2022 onwards).
- day-of-the-week: indicator variables to account for day-of-the-week reporting effects in the numbers of cases.
- urban/rural status: binary covariates for whether the NUTS-3 region *i* is classified as urban, intermediate urban, intermediate rural or rural.
- seasonality: sinusoidal terms with annual periodicity to account for their effects on local transmission. The amplitude and offset of the seasonality function are estimated by the model.

The covariates above cover the impact of local immunity (due to vaccine or previous infection), testing patterns, variants and seasonality on the risks of reported cases. We Included covariates that quantify the association between number of tests and incidence so that the model may capture changes in surveillance and reporting patterns, whereby drops in testing can be associated with changes in the number of new cases. For instance, in age groups with a lower proportion of hospitalised cases (e.g. younger age groups) the reporting rate may vary greatly if only severe cases are tested and reported. The covariates quantifying the association between infection or vaccine-acquired immunity and transmission risk depend on various thresholds (e.g. recent incidence corresponds to cases reported in the last month; the second vaccine dose is only taken into account during 120 days). These thresholds can easily be changed in the code to generate sensitivity analyses, or to adapt to the characteristics of new variants or vaccines.

### Model fitting

We fit the model to case data reported between September 2020 and the latest reported date for each country (currently end of April 2023) to estimate the regression coefficients *β* ^*(ϕ)*^ and *β*^*(v)*^. The model is fit separately for each country. We do this via maximum-likelihood estimation, as implemented in the *surveillance* and *hhh4addon* R packages.

### Case forecasts

We use the parameter estimates from the model fitting to generate four-week-ahead forecasts of daily case numbers in each region and age group by simulating the model forward 28 days with projected values of the covariates. We used the latest value of the covariates describing vaccine coverage and testing, and the mean value over the past 30 days for the number of cases in the rest of Europe in the past month. We run 500 simulations for each country to account for stochasticity and parameter uncertainty, and output the median, 2.5th, 25th, 75th, and 97.5th percentiles of the predicted distribution for each date and region (and age group for the age-stratified model) for visualisation in the RShiny app.

### Death forecasts

We use simple linear regression models to generate four-week-ahead forecasts of weekly numbers of reported deaths. To do so, we estimate the Case Fatality Rate (CFR), and combine it with the recent number of reported cases to forecast the number of deaths, We first aggregate the number of reported cases and deaths by week, and compute a proxy for the age-stratified CFR assuming a three-week delay between weekly reported cases and reported deaths (42– 44). Specifically, we compute the CFR for age group *ai* in region *i* at week *w* as:

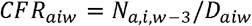

where *N*_*aiw*_ and *D*_*aiw*_ are the numbers of cases and deaths in age group *a*, in NUTS-2 region *i*, in week w. The death forecast model was implemented at a NUTS-2 geographical scale since death data was not always available at a NUTS-3 level.

We then calculate *ΔCFR*_*aiwx*_, the change in CFR, and *ΔN*_*aiwx*_, the change in the number of cases, between the prediction date and the forecast horizon:

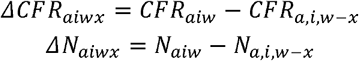

where *x* is the forecast horizon (1, 2, or 3 weeks). We implement a linear regression model for each forecast horizon and age group, with *ΔCFR* as outcome, and *ΔN* as explanatory variable, again using a three-week delay:

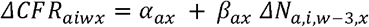

We then use the estimates of *α*_*ax*_ and *β*_*ax*_ to predict *ΔCFR* between the last week of reported case data (*w*_*pred*_) and the forecast dates (*w*_*pred*_ + 1, *w*_*pred*_ + 2, *w*_*pred*_ + 3), and calculate the predicted CFR as:

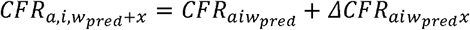

(using a sample of 10 values of *ΔCFR*_*aiwx*_, computed from the mean estimate and the prediction interval of the linear regression). Finally, we draw the number of new weekly deaths each week using a binomial distribution, from the estimated CFR, and the number of cases reported three weeks before:

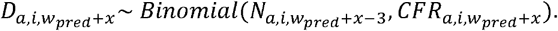

We draw 500 forecasts for 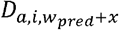 in total (50 per value of 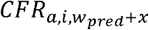). Given the three-week delay between cases and deaths, one-, two-, and three-week ahead death forecasts are generated using already reported case data. In contrast, four-week-ahead death forecasts require one-week-ahead case forecasts. We estimate the change in CFR using the regression parameters from the three-week-ahead forecasts and the four-week change in number of cases in the one-week-ahead case forecasts

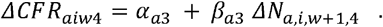

### Scenario simulations

To explore the impact of variations in transmission intensity or implementation of non-pharmaceutical interventions (NPIs), we generate 28-day forecasts under combinations of the following scenarios, by changing the values of the epidemic predictor *ϕ*_*ait*_, or the endemic predictor *v*_*ait*_ :

- Moderate (20%) or large (40%) increase in transmission intensity (*ϕ*_*ait*_), due to inherent properties of the pathogen (i.e. emergence of a new, more transmissible variant), or to changes in behaviour.
- Moderate (20%) or large (40%) decrease in transmission intensity (*ϕ*_*ait*_), due to changes in human behaviour or NPIs. Furthermore, for countries where an age-stratified model was implemented, this decrease can be targeted at a certain age group (children and teenagers below 20 years old, adults between 20 and 60 years old, or older inhabitants), and implemented one or two weeks after the current date.
- Removal of all importations, for instance due to border closure (i.e. *v*_*ait*_ = 0).

All scenarios affect every region in the same way (i.e. we do not consider local NPIs). We generate 100 simulations under each scenario.

## Data

Several publicly available data sources are used to implement the model and are summarised in Table 1. Since compilation of COVID-19 surveillance reports in centralised databases was interrupted in early 2022 in many countries, the majority of the data, including the case and death data, is imported from country-specific sources.

**Table 1:** Case, death, vaccination, test and population data sources used in each country and their spatial and temporal resolution and age stratification.

| Country | Metropolitan France | Czechia | Italy |
| --- | --- | --- | --- |
| Case data source | Santé Publique France (**daily age-stratified** number of cases in each **NUTS-3 region** (48)) | Ministerstvo Zdravotnictví České Republiky (**daily age-stratified** number of cases in each **NUTS-3 region** (49)) | Dipartimento Della Protezione Civile (**daily non- age-stratified** number of cases reported per **NUTS-3 region** (50)) |
| Death data source | Santé Publique France (**daily age-stratified** number of deaths in each **NUTS-1 region** (48)) | Ministerstvo Zdravotnictví České Republiky (**daily age-stratified** number of deaths in each **NUTS-3 region** (49)) | Dipartimento Della Protezione Civile (**daily non- age-stratified** number of deaths reported per **NUTS-2 region** (50)) |
| Vaccination data source | l'Assurance Maladie (**weekly age-stratified** number of doses in each **NUTS-3 region** (51)) | Ministerstvo Zdravotnictví České Republiky (**daily age-stratified** number of doses in each **NUTS-3 region** (49)) | ECDC vaccination database (**overall weekly** number of doses administered in each **NUTS-2 region** (52)) |
| Test data source | Santé Publique France (**daily age-stratified** number of tests in each **NUTS-3 region** (48)) | Ministerstvo Zdravotnictví České Republiky (**daily national age-stratified** number of tests (49)) | Dipartimento Della Protezione Civile (**weekly national** number of tests (50)) |
| Population data source | INSEE (Number of inhabitants per **NUTS-3 region and age group** (53)) | Eurostat (Number of inhabitants per **NUTS-3 region and age group** (54)) | Google COVID-19 Open Data (Number of inhabitants per **NUTS-3 region** (55)) |

### Case and death data

Local case data at a NUTS-3 level was used in all countries (age-stratified in France and Czechia). In Italy and France, death data was only available at NUTS-1/NUTS-2 level.

### Covariate data

Where available, daily age-stratified (for the age-stratified model) vaccination data at NUTS-3 regional level is used, though most sources provide weekly data, and a two-week delay for protection from each dose to develop is assumed. Publicly available testing data varies considerably in spatial resolution (for some countries only national data is available) and age stratification (for some countries only total data is available), so age-stratified national testing data is used as a default and subnational data used when available. Age-stratified regional population data is drawn from a different source for each country (Table 1). Data on the proportion of different variants among sequenced cases is drawn from the ECDC variant database (45). The urban-rural status of each NUTS-3 region is taken from the Eurostat database (database labelled “urban-rural remoteness”) (46). Daily numbers of cases in the rest of Europe are taken from the World Health Organisation database of daily numbers of cases and deaths in different countries (47).

### Calibration analysis

We evaluate the ability of the model to generate accurate and reliable case and death forecasts in each country by fitting the model up to a set of dates in a calibration period, generating 28-day-ahead forecasts from these dates using the model fits, and comparing these forecasts with the data. The forecasts are generated by re-fitting the model for each calibration date (i.e. we do not use the data posterior to the calibration date to generate the forecasts). We used weekly dates from the last six months of data as calibration period (29th October 2022 to 22nd April 2023, (i.e. 25 calibration dates). A model is deemed well-calibrated if it can identify its own uncertainty in making predictions, i.e. if the data points are evenly distributed across the prediction intervals generated by the model. Since the reliability of our forecasts is likely to depend on the prediction horizon, we compare the performance of the model for one-, two-, three-, and four-week-ahead forecasts, to determine at which point the quality of the forecasts declines.

We use various metrics and figures to evaluate the calibration. Firstly, we visually compare our weekly national-level case and death forecasts, obtained by summing up the age-stratified and local forecasts, with the ensemble forecasts produced by the European COVID-19 Forecast Hub, which serves as a benchmark of short-term forecasts of COVID-19 incidence. We also compare our national-level forecasts with the ensemble forecasts quantitatively via the weighted interval score (WIS) (31), a proper scoring rule (i.e. one that measures both calibration – how accurate the forecasts are – and sharpness – how precise the forecasts are) for quantile forecasts, and the squared error of the median. This comparison assesses whether the overall performance of our model is in line with other COVID-19 prediction models.

Secondly, we evaluate our local forecasts against those of a baseline model (chosen as the Endemic-Epidemic model without transmission between regions, covariates or seasonality, i.e. the model in Equations (1)-(4) but with *w*_*ji*_ =1 when *j*=i and *w*_*ji*_ =0 otherwise and *log*(*ϕ*_*it*_)= *α*^*(ϕ)*^ and *log*(*v*_*it*_)= *α*^*(v)*^, using proper scoring rules, namely the ranked probability score (RPS), Dawid-Sebastiani score (DSS) and squared error score (SES) (see Supplementary Material for definitions). We also generate the predictive probability distribution of the local, age-stratified (if available) forecasts at each calibration date. We then use the Probability Integral Transform (PIT) histogram to assess the calibration of the model: in models with good calibration, the data should follow the predictive probability distribution, and the PIT histogram should be uniform. We computed a non-randomised yet uniform version of the PIT histogram, to correct for the use of discrete values, as described in Czado et al (56).

### Comparison of age-stratified and non-age-stratified model

As age-stratified case and death data is only available for certain countries, we explore the impact of fitting the model to subnational total case counts for France and Czechia on the ability of the model to predict subnational total numbers of cases and deaths. We do this by fitting the non-age-stratified equivalent of the age-stratified model in Equations (3)-(4) and (A1)-(A2) with the age-stratified covariates removed, i.e. Equations (1)-(2) and (A4) (see Supplementary Material), and re-running the calibration analysis of the predictive performance of the model described above.

### RShiny Application

All forecasts and predictors generated by the model are available in an R Shiny Application (https://github.com/EU-ECDC/RShinyCovidApp). The users can use the application to see the latest case and death forecasts of the model in each country, compare past forecasts to recent data points, generate different transmission and NPI scenarios, and observe the regions most at risk of transmission according to the model. The forecasts contained in the application are updated weekly.

## Results

### Calibration analysis: assessing the performance of the model

#### Case forecasts calibration

Firstly, we compute the weekly case forecasts generated by the model across all regions and age groups, by summing up all local age stratified forecasts, across the calibration period (last six months of data). We then compare the one, two, three, and four-week ahead forecasts with the observed data, and the weekly forecasts from the European COVID-19 Forecast Hub.

The comparison of the forecasts is shown in Figure 1 and Table 2. It demonstrates that our model was able to reliably capture the dynamics observed in the data up to two weeks ahead. The magnitude of the peak of the different outbreaks is accurately estimated in all three countries, and the trend forecasted by our model corresponds to the data. Beyond a two-week forecast horizon, the difference between the forecasts and the data becomes starker, in particular during outbreaks (for instance around December 2022 in France), with changes in case numbers being captured with a bigger lag in the forecasts. Based on the squared error of the median, the median forecasts generated by the model are on par with the ensemble forecasts from the European COVID-19 Forecast Hub across all forecast horizons (Table 2). There are periods where our model outperforms the ensemble model (e.g. two-week ahead forecasts between September and December 2022 in Italy), and others where the ensemble model outperforms ours (January-March 2023 in Czechia across all forecast horizons).

**Figure 1:**
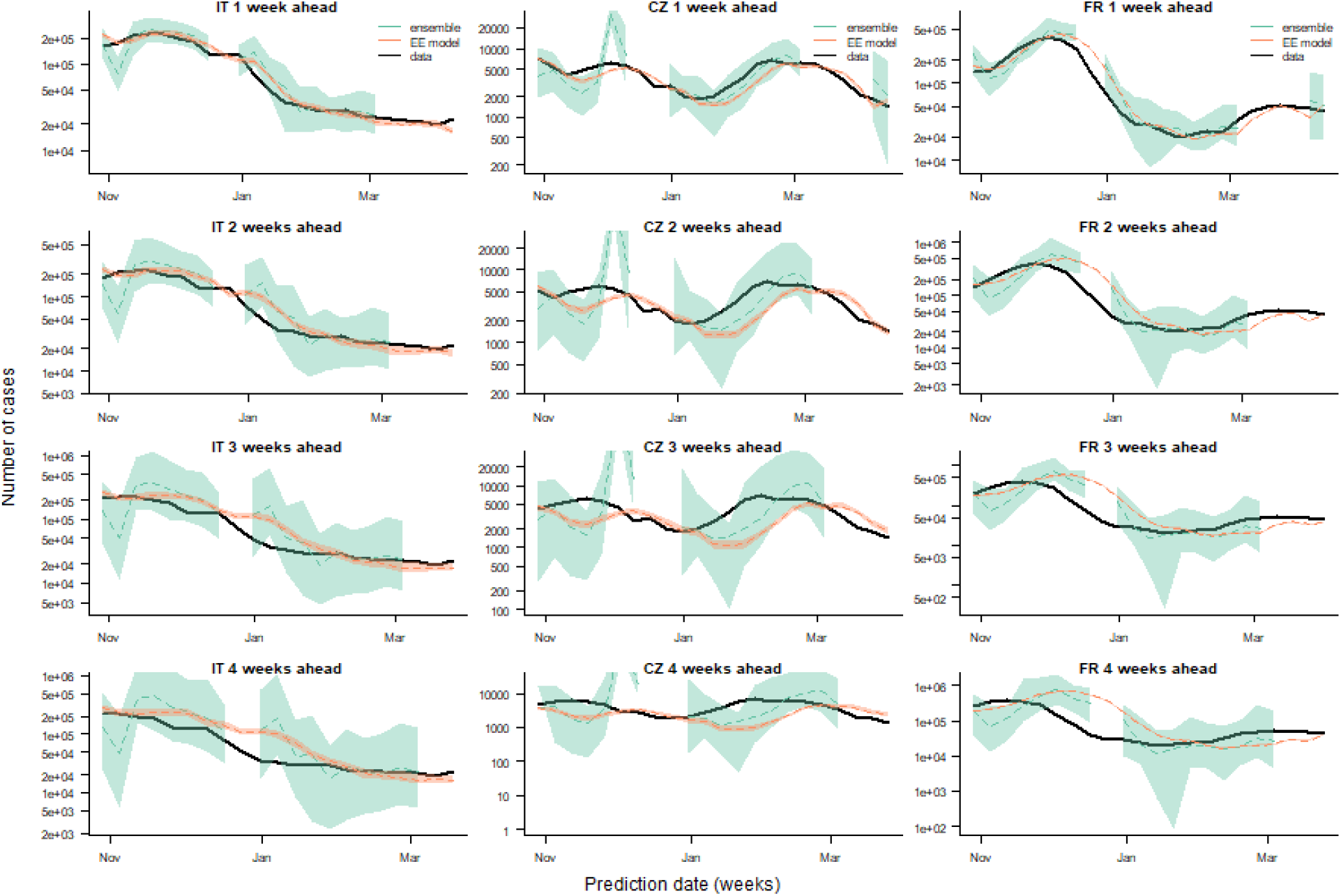
Comparison of one-, two-, three- and four-week-ahead national-level case forecasts (rows, top to bottom) for Italy, Czechia and France (columns, left to right) from our model (Endemic-Epidemic (EE) model) with ensemble forecasts from the European COVID-19 Forecast Hub (ensemble) and observed cases (data) for a calibration period from 29 October 2022 to 22 April 2023. Dashed lines show median forecasts, shaded regions the 95% prediction interval for the forecasts. The weekly number of cases is shown using a logged axis.

**Table 2:**
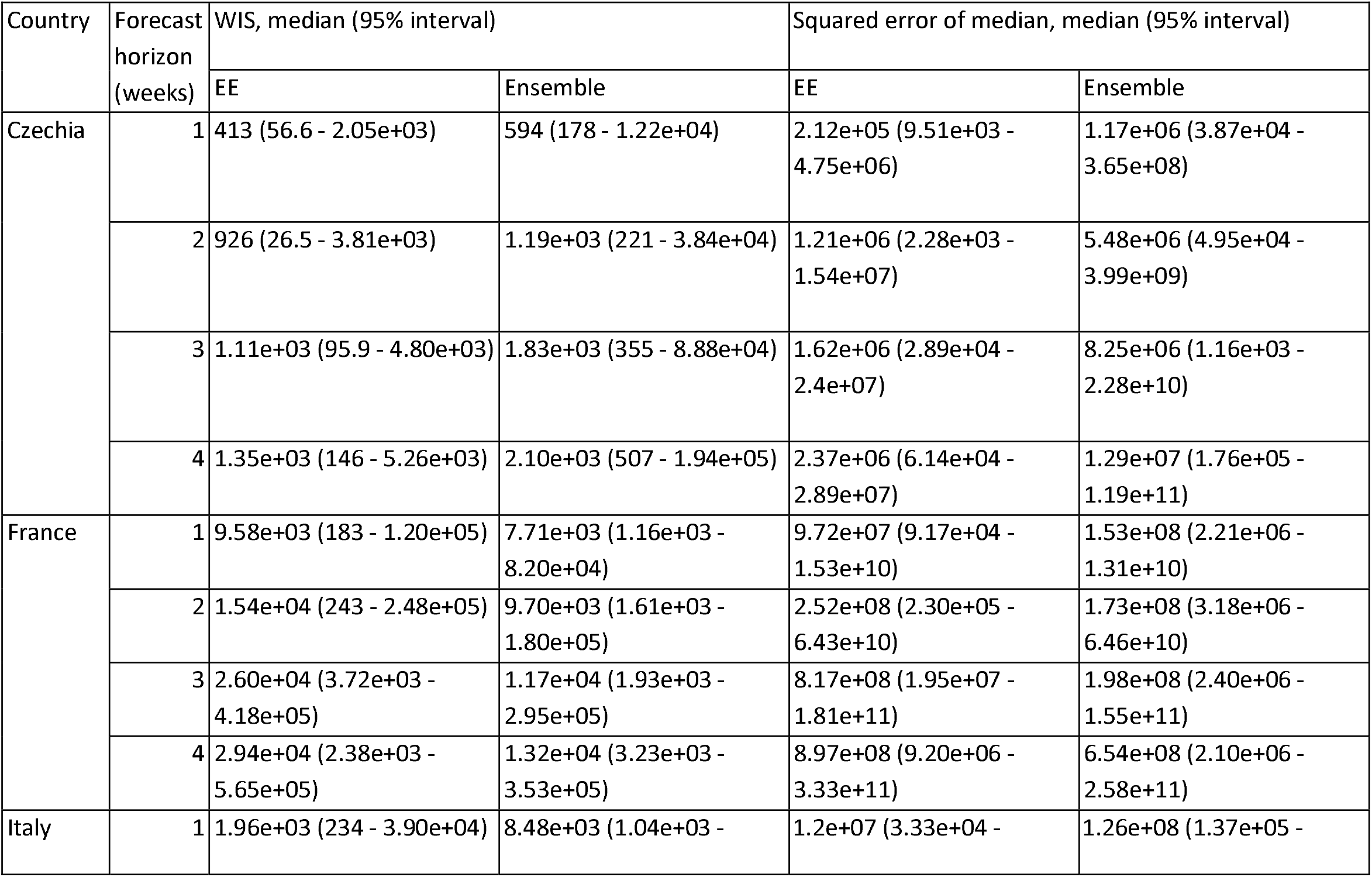
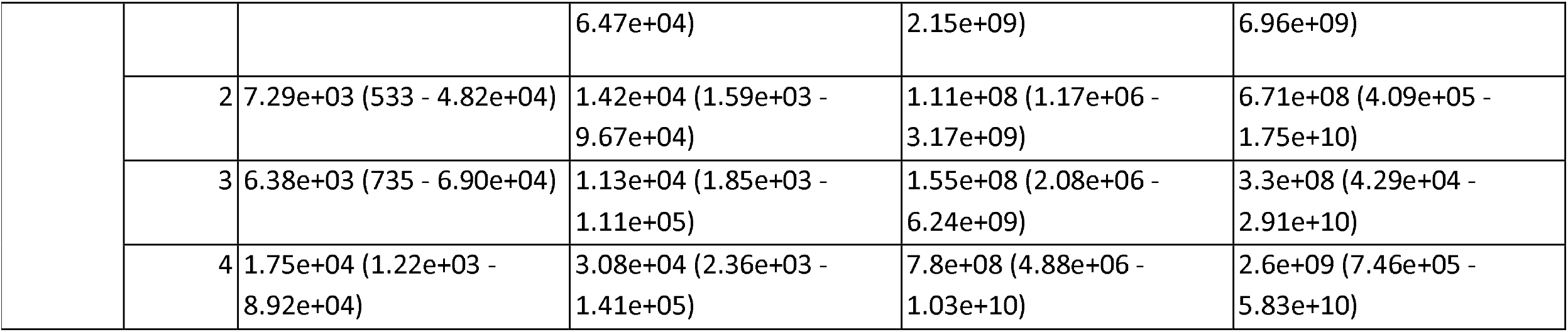
Median and 95% interval of weighted interval score (WIS) and squared error of the median of one-, two-, three- and four-week-ahead national-level case forecasts for Czechia, France and Italy for our model (Endemic-Epidemic, EE) and European COVID-19 Forecast Hub ensemble model (Ensemble) across all time points in the prediction period from 29 October 2022 to 22 April 2023.

| Country | Forecast horizon (weeks) | WIS, median (95% interval) |  | Squared error of median, median (95% interval) |  |
| --- | --- | --- | --- | --- | --- |
|  |  | EE | Ensemble | EE | Ensemble |
| Czechia | 1 | 413 (56.6 - 2.05e+03) | 594 (178 - 1.22e+04) | 2.12e+05 (9.51e+03 - 4.75e+06) | 1.17e+06 (3.87e+04 - 3.65e+08) |
|  | 2 | 926 (26.5 - 3.81e+03) | 1.19e+03 (221 - 3.84e+04) | 1.21e+06 (2.28e+03 - 1.54e+07) | 5.48e+06 (4.95e+04 - 3.99e+09) |
|  | 3 | 1.11e+03 (95.9 - 4.80e+03) | 1.83e+03 (355 - 8.88e+04) | 1.62e+06 (2.89e+04 - 2.4e+07) | 8.25e+06 (1.16e+03 - 2.28e+10) |
|  | 4 | 1.35e+03 (146 - 5.26e+03) | 2.10e+03 (507 - 1.94e+05) | 2.37e+06 (6.14e+04 - 2.89e+07) | 1.29e+07 (1.76e+05 - 1.19e+11) |
| France | 1 | 9.58e+03 (183 - 1.20e+05) | 7.71e+03 (1.16e+03 - 8.20e+04) | 9.72e+07 (9.17e+04 - 1.53e+10) | 1.53e+08 (2.21e+06 - 1.31e+10) |
|  | 2 | 1.54e+04 (243 - 2.48e+05) | 9.70e+03 (1.61e+03 - 1.80e+05) | 2.52e+08 (2.30e+05 - 6.43e+10) | 1.73e+08 (3.18e+06 - 6.46e+10) |
|  | 3 | 2.60e+04 (3.72e+03 - 4.18e+05) | 1.17e+04 (1.93e+03 - 2.95e+05) | 8.17e+08 (1.95e+07 - 1.81e+11) | 1.98e+08 (2.40e+06 - 1.55e+11) |
|  | 4 | 2.94e+04 (2.38e+03 - 5.65e+05) | 1.32e+04 (3.23e+03 - 3.53e+05) | 8.97e+08 (9.20e+06 - 3.33e+11) | 6.54e+08 (2.10e+06 - 2.58e+11) |
| Italy | 1 | 1.96e+03 (234 - 3.90e+04) | 8.48e+03 (1.04e+03 - | 1.2e+07 (3.33e+04 - | 1.26e+08 (1.37e+05 - |
|  |  |  | 6.47e+04) | 2.15e+09) | 6.96e+09) |
|  | 2 | 7.29e+03 (533 - 4.82e+04) | 1.42e+04 (1.59e+03 - 9.67e+04) | 1.11e+08 (1.17e+06 - 3.17e+09) | 6.71e+08 (4.09e+05 - 1.75e+10) |
|  | 3 | 6.38e+03 (735 - 6.90e+04) | 1.13e+04 (1.85e+03 - 1.11e+05) | 1.55e+08 (2.08e+06 - 6.24e+09) | 3.3e+08 (4.29e+04 - 2.91e+10) |
|  | 4 | 1.75e+04 (1.22e+03 - 8.92e+04) | 3.08e+04 (2.36e+03 - 1.41e+05) | 7.8e+08 (4.88e+06 - 1.03e+10) | 2.6e+09 (7.46e+05 - 5.83e+10) |

However, the dispersion of our forecasts is considerably narrower than that of the ensemble forecasts, particularly for France. The data points are therefore more often outside the 95% prediction interval of our forecasts, despite the median forecasts being close to the data points, and the median and 95% interval of the weighted interval score is higher for France for our model than the ensemble model (Table 2). The narrow prediction interval is a consequence of aggregating all local and age-stratified forecasts to get the national-level estimate rather than directly estimating the national number of new cases: since the dispersion of a sum of negative binomial random variables is much smaller than that of each individual negative binomial random variable. This is also why the prediction interval is narrowest in France, where the number of groups is highest (846 groups = 94 regions times 9 age groups).

We now assess whether the dispersion of local estimates is in line with the data (i.e. whether the prediction intervals from the model include the observed data points). To do so, we compute the case PIT histogram for each country and forecast horizon (Figure 2). We also generate PIT histograms stratified by broad age groups for France and Czechia (Supplementary Figures 2 and 3). In one-week-ahead forecasts, we observe an inverse U-shaped histogram in Italy, indicating that the forecasts are slightly overdispersed. The performance of the age-stratified model varies in the different age groups. The model tends to overestimate the number of cases in younger age groups (Supplementary Figures 2 and 3), while the histograms in age groups between 20 and 80 years old were flat, indicating good calibration. The two-week ahead PIT histogram for Italy also shows very good performance, with little sign of bias, although the model tends to slightly overestimate the number of cases (few observations fall into the highest categories). Over longer time horizons, the PIT histogram becomes U-shaped, i.e. skewed towards extreme values, showing that the forecasts generated by the model are too confident.

**Figure 2:**
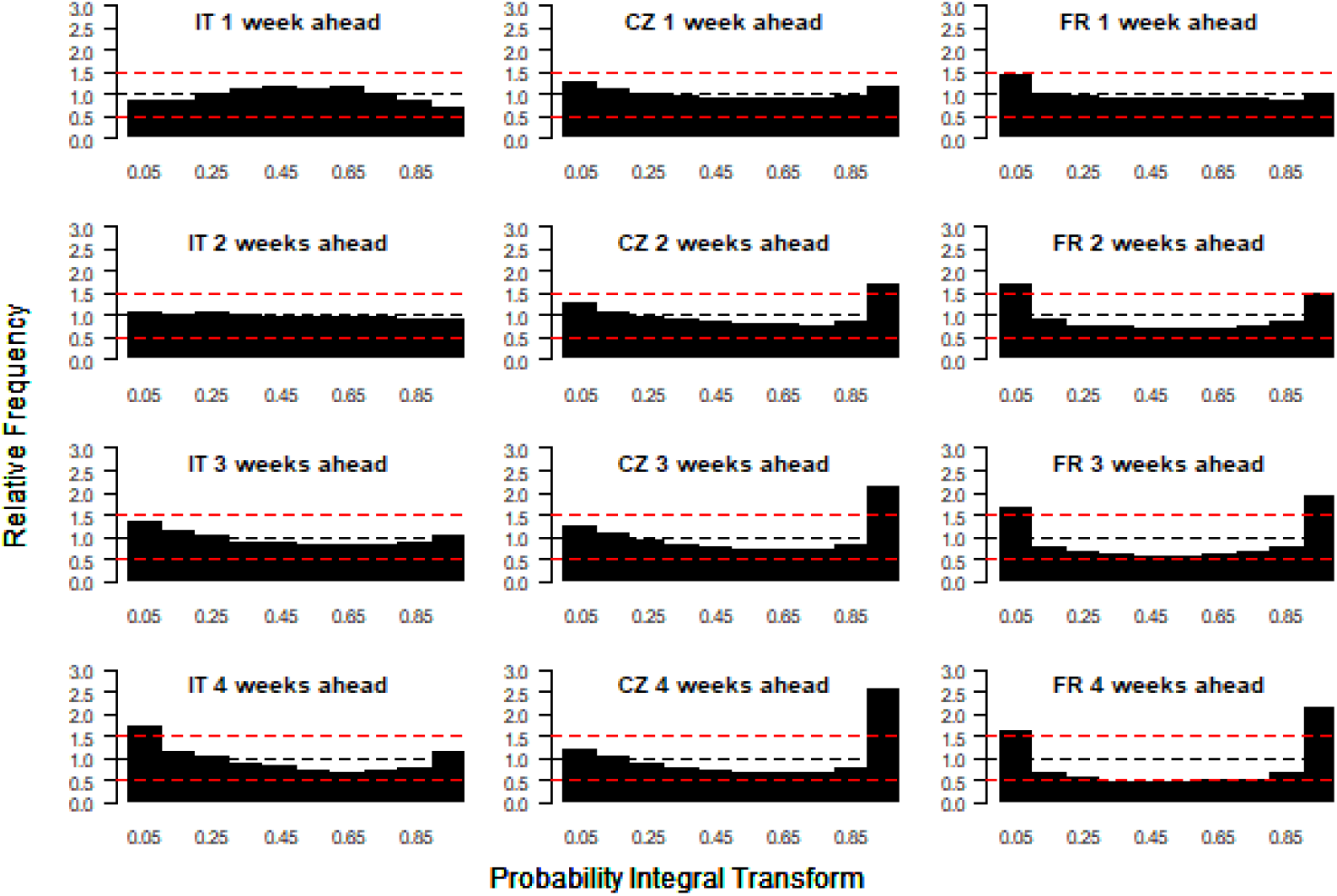
Probability integral transform histograms showing the calibration of the daily case forecasts from the model for Italy, Czechia and France (columns, left to right) for one-, two-, three- and four-week-ahead forecast horizons (rows, top to bottom) for 29 October 2022 to 22 April 2023. Uniform histograms indicate well-calibrated forecasts, while U- and inverse U-shaped histograms indicate underdispersed and overdispersed forecasts respectively. Red dashed lines at relative frequencies of 0.5 and 1.5 show reasonable bounds for calibration compared to desired relative frequency of 1 (blue dashed line).

**Figure 3:**
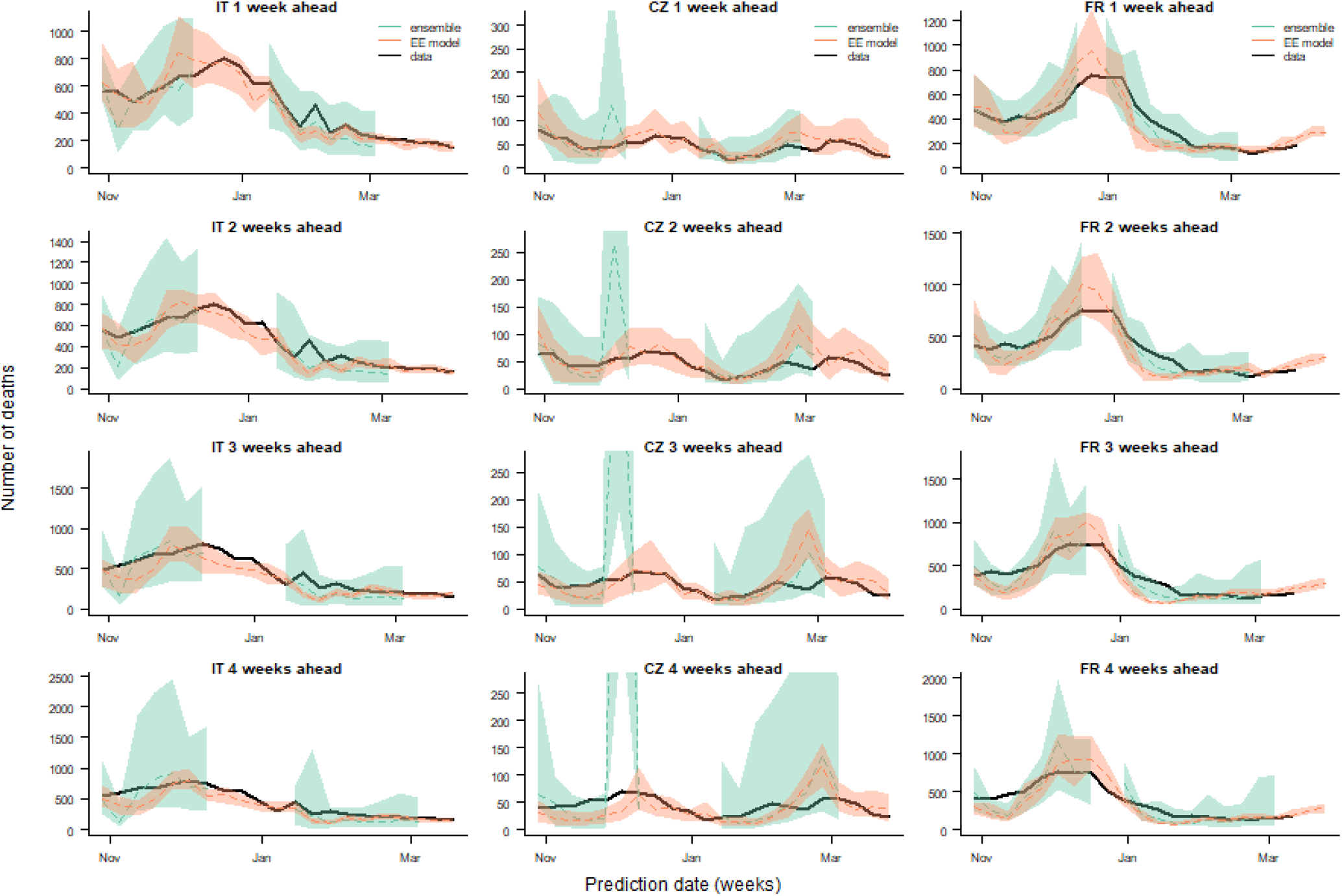
Comparison of one-, two-, three- and four-week-ahead death forecasts (rows, top to bottom) for Italy, Czechia and France (columns, left to right) from our model (Endemic-Epidemic (EE) model) with ensemble forecasts from the European COVID-19 Forecast Hub (ensemble) and observed deaths (data) for a calibration period from 29 October 2022 to 22 April 2023. Dashed lines show median forecasts, shaded regions the 95% prediction interval for the forecasts.

The histograms for Czechia and France are U-shaped, indicating that the model is overly confident (i.e. the prediction interval is too narrow). The overestimation of cases in younger age groups may be due to case incidence being determined by the age-structured contact rates used in the model (derived from contact surveys), but cases in this age group may also be under-reported as they typically have milder symptoms, and so are harder to spot without active case finding campaigns. Forecasts past two weeks ahead are more strongly biased, and underdispersed, but better performance is still observed for Italy, where the model is not age-stratified. Similar underdispersion was observed in Czechia and France when a non-age-stratified model was implemented (Supplementary Figures 6 and 7).

We also generated local forecasts using a baseline model (with no transmission between regions, covariates, seasonality, or age-specific intercepts), and compared them to the forecasts from the full model (Supplementary Table 2). The addition of transmission between regions, covariates and seasonality to the model substantially improved the predictive performance up to 2 weeks ahead for all countries, with an average 38% improvement across the three countries in the median RPS at a 2-week forecast horizon. This was especially true in Czechia and France, where the age-specific dynamics were hard to capture without covariates.

#### Death forecasts calibration

The death forecast comparison is shown in Figure 3 and Table 3. The performance of the model is not homogeneous across countries. The model performs better in Italy and Czechia, where the median estimate is closer to the observed data, and the 95% prediction interval includes the data more often than in France. In France, the model was not able to accurately capture the drop in number of deaths following the outbreak in December 2022. For most dates, the median estimate of the number of deaths is similar in our model and the European COVID-19 Forecast Hub ensemble model (Figure 3), and the median of the squared errors of the median predictions across all time points in the prediction period is similar (Table 3). The predictive accuracy and coverage of our model is closer to that of the ensemble model for the death forecasts than the case forecasts (the WIS is closer, Table 3), and the impact of forecast horizon on the performance is not as severe as for the case forecasts, for instance for Italy four-week-ahead forecasts are still in line with the number of reported deaths (Figure 3). This is due to the larger prediction intervals generated with the death model relative to the number of deaths.

**Table 3:**
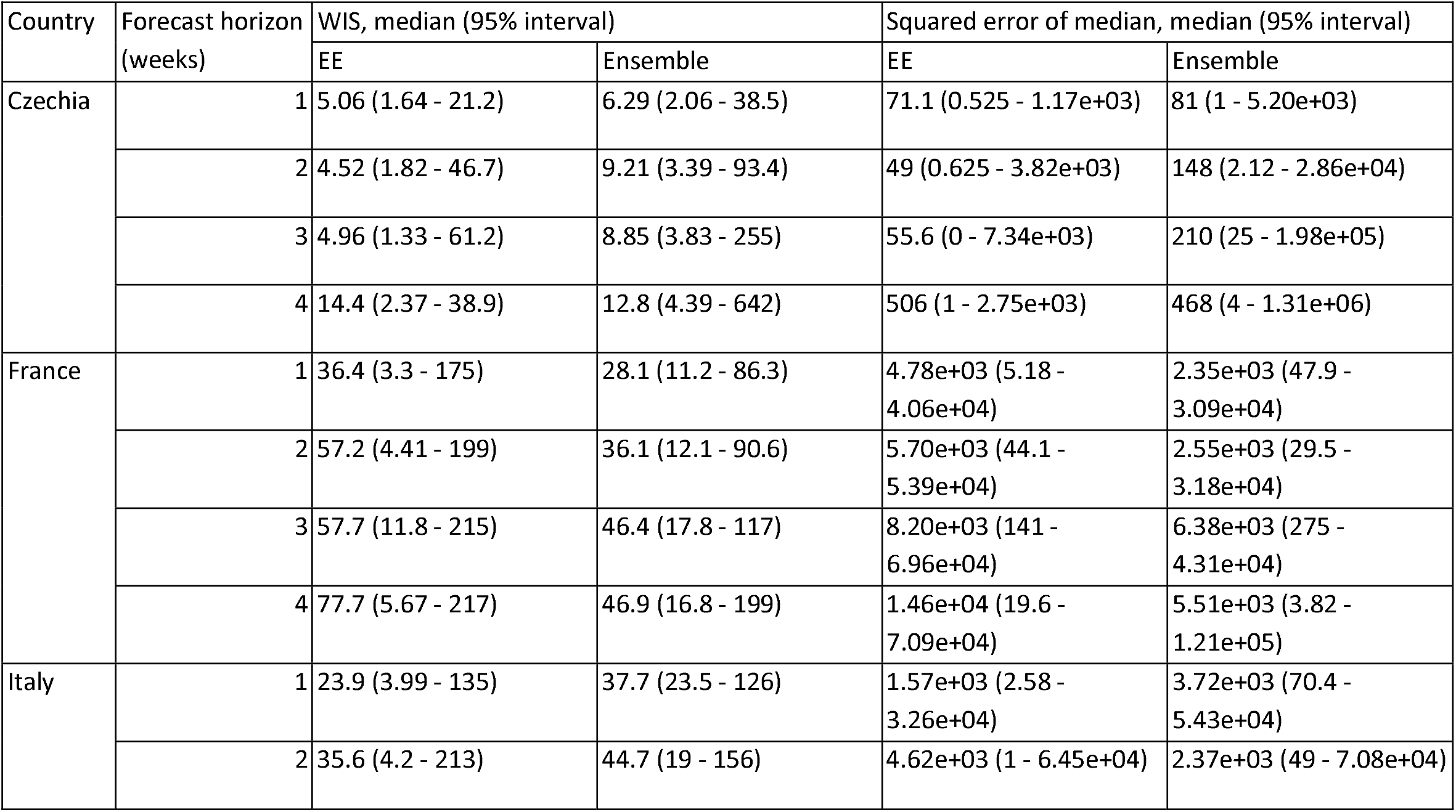
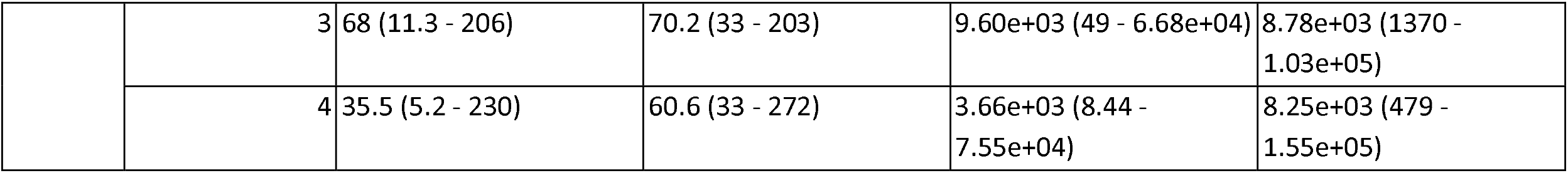
Median and 95% interval of weighted interval score (WIS) and squared error of the median of one-, two-, three- and four-week-ahead national-level death forecasts for Czechia, France and Italy for our model (Endemic-Epidemic, EE) and European COVID-19 Forecast Hub ensemble model (ensemble) across all time points in the prediction period from 29 October 2022 to 22 April 2023.

| Country | Forecast horizon (weeks) | WIS, median (95% interval) |  | Squared error of median, median (95% interval) |  |
| --- | --- | --- | --- | --- | --- |
|  |  | EE | Ensemble | EE | Ensemble |
| Czechia | 1 | 5.06 (1.64 - 21.2) | 6.29 (2.06 - 38.5) | 71.1 (0.525 - 1.17e+03) | 81 (1 - 5.20e+03) |
|  | 2 | 4.52 (1.82 - 46.7) | 9.21 (3.39 - 93.4) | 49 (0.625 - 3.82e+03) | 148 (2.12 - 2.86e+04) |
|  | 3 | 4.96 (1.33 - 61.2) | 8.85 (3.83 - 255) | 55.6 (0 - 7.34e+03) | 210 (25 - 1.98e+05) |
|  | 4 | 14.4 (2.37 - 38.9) | 12.8 (4.39 - 642) | 506 (1 - 2.75e+03) | 468 (4 - 1.31e+06) |
| France | 1 | 36.4 (3.3 - 175) | 28.1 (11.2 - 86.3) | 4.78e+03 (5.18 - 4.06e+04) | 2.35e+03 (47.9 - 3.09e+04) |
|  | 2 | 57.2 (4.41 - 199) | 36.1 (12.1 - 90.6) | 5.70e+03 (44.1 - 5.39e+04) | 2.55e+03 (29.5 - 3.18e+04) |
|  | 3 | 57.7 (11.8 - 215) | 46.4 (17.8 - 117) | 8.20e+03 (141 - 6.96e+04) | 6.38e+03 (275 - 4.31e+04) |
|  | 4 | 77.7 (5.67 - 217) | 46.9 (16.8 - 199) | 1.46e+04 (19.6 - 7.09e+04) | 5.51e+03 (3.82 - 1.21e+05) |
| Italy | 1 | 23.9 (3.99 - 135) | 37.7 (23.5 - 126) | 1.57e+03 (2.58 - 3.26e+04) | 3.72e+03 (70.4 - 5.43e+04) |
|  | 2 | 35.6 (4.2 - 213) | 44.7 (19 - 156) | 4.62e+03 (1 - 6.45e+04) | 2.37e+03 (49 - 7.08e+04) |
|  | 3 | 68 (11.3 - 206) | 70.2 (33 - 203) | 9.60e+03 (49 - 6.68e+04) | 8.78e+03 (1370 - 1.03e+05) |
|  | 4 | 35.5 (5.2 - 230) | 60.6 (33 - 272) | 3.66e+03 (8.44 - 7.55e+04) | 8.25e+03 (479 - 1.55e+05) |

For Czechia and France, the PIT histograms generated for the local age-stratified weekly death forecasts follow much more uniform distributions than those for the case forecasts, for all forecast horizons (Figure 4). Since the model was age stratified for both countries, the majority of weekly death counts in the data were 0, which was easier for the model to capture and forecast. In Italy, the upper bound of the prediction interval tends to be too low, since few observed weekly death counts fall into the lower categories of the PIT histograms. Therefore, our model tends to underestimate the number of weekly deaths per region in Italy. The age-stratified PIT histograms in Czechia and France (Supplementary Figures 4 and 5) are uniform for younger age groups (where almost all observations are 0), and follow an inverse U-shape in older age groups, indicating that the forecasts are slightly overdispersed.

**Figure 4:**
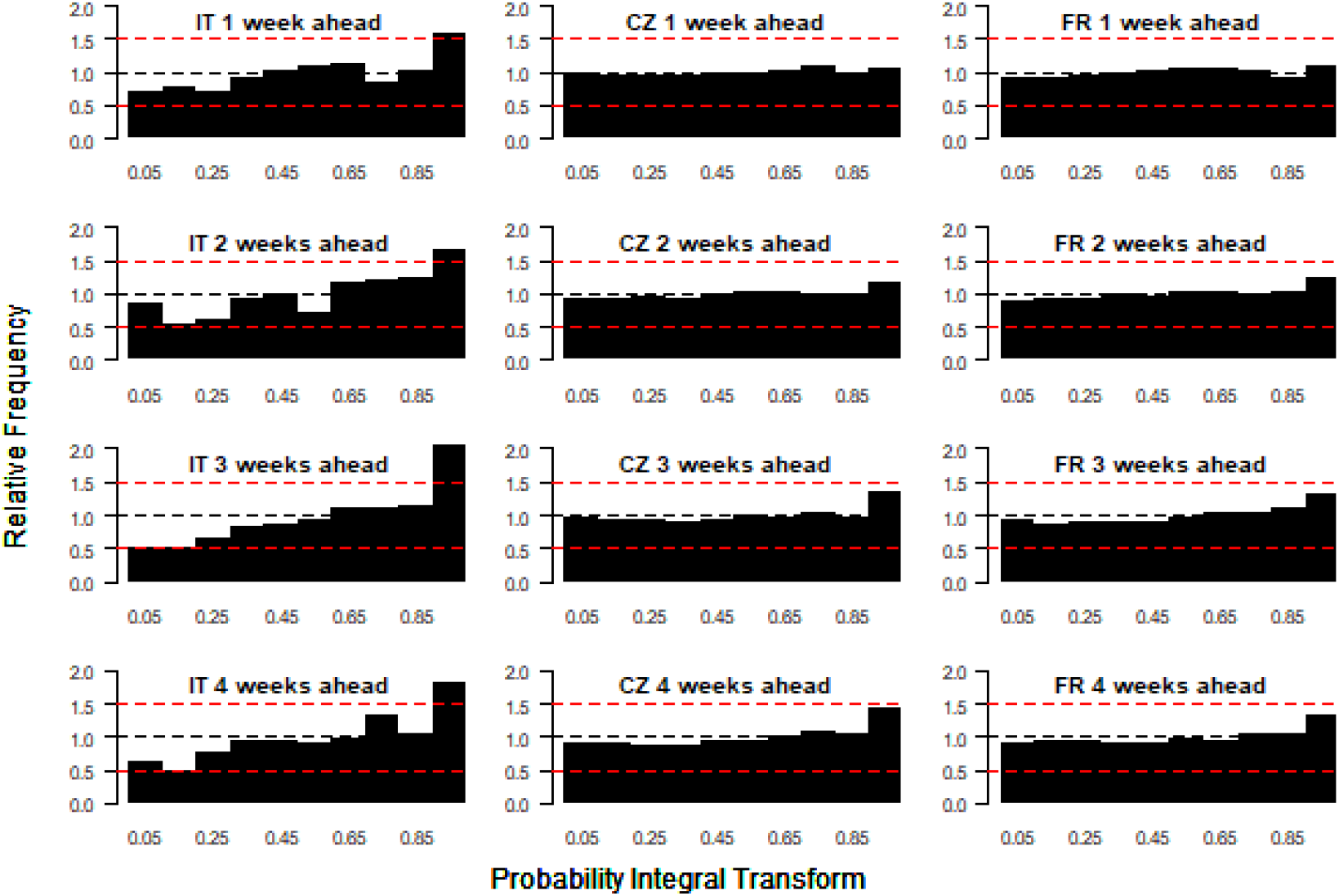
Probability integral transform histograms showing the calibration of the weekly death forecasts from the model for Italy, Czechia and France (columns, left to right) for one-, two-, three- and four-week-ahead forecast horizons (rows, top to bottom) for 29 October 2022 to 22 April 2023. Red dashed lines at relative frequencies of 0.5 and 1.5 show reasonable bounds for calibration compared to desired relative frequency of 1 (blue dashed line).

**Figure 5:**
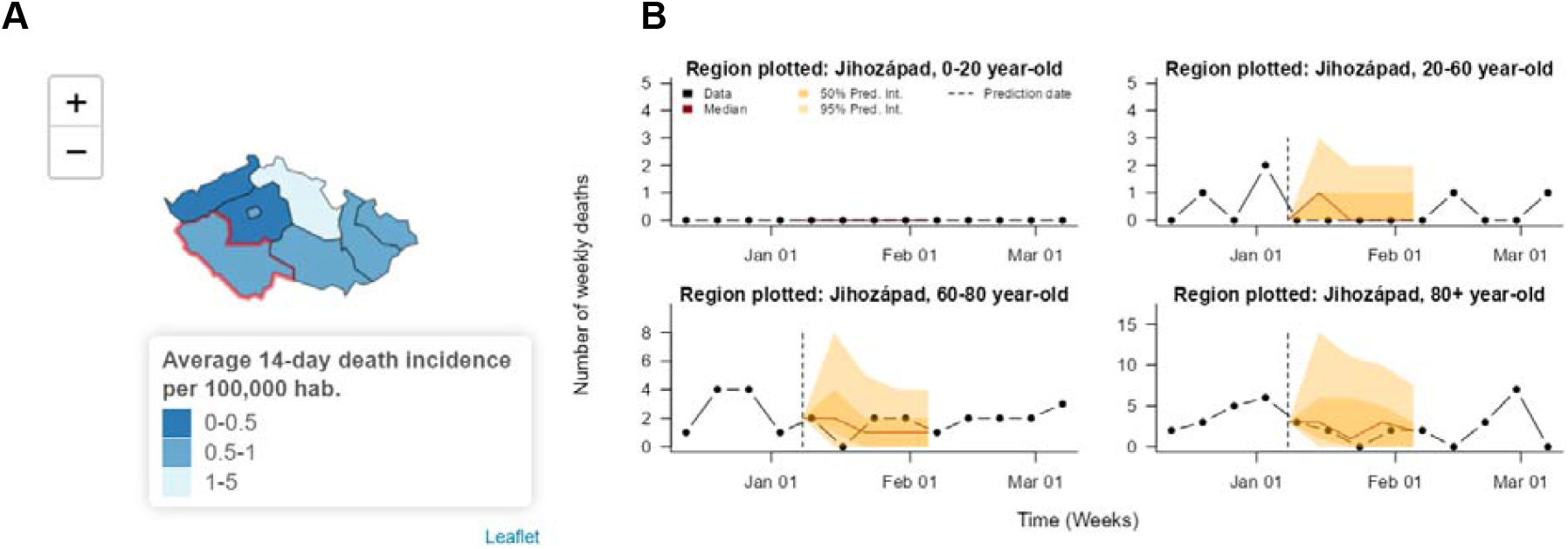
Visualisation of COVID-19 death forecasts for a region of Czechia in the RShiny app. (A) Map of forecasted 14-day incidence of cases per 100,000 inhabitants from 9th to 23rd January 2023 at NUTS-3 level. (B) Age-stratified 28-day-ahead case forecasts for the region outlined in red in (A). Since the prediction date is 4 weeks in the past, the observed number of deaths is also plotted to allow assessment of the accuracy of the forecasts. The closeness of the median forecast to the observed data and the fact that the 95% prediction intervals cover nearly all of the observed data points indicate the good predictive performance of the model for this region. Time series forecast plots for other regions can be viewed in the app by clicking on those regions in the map. Together, panels (A) and (B) can be used to identify which regions appear to be at risk of higher burden.

### Visualising the predictions in the R Shiny Application

The RShiny app is designed to make three features accessible to the user:

1. Forecasts: current and previous four-week ahead forecasts of the number of cases and deaths in each region (and age group when available)
2. Predictors: The risks of secondary transmission and importations estimated by the model
3. Scenarios: The impact of changes in transmission, be it due to more infectious variants, changes in behaviour, or NPIs on case and death forecasts. These changes in transmission represent the potential impact of control measures on transmission, and should be interpreted with caution in a constantly changing epidemiological situation (impacted for example by behaviour, adherence and other factors). Similarly, this model only considers the epidemiological impact of NPIs, the social or economic costs of different control measures is not considered in this analysis.

#### Previous and current forecasts

Case and death forecasts can be viewed via a map of the predicted median case/death incidence over the next two weeks, or via time series plots by region (and age group when age-stratified data is available) of the median numbers of daily cases/weekly deaths over the next four weeks (with 50% and 95% prediction intervals) (Figure 5). Forecasts can be visualised at different geographical scales, either at NUTS-3 level or NUTS-1/NUTS-2 level. The accuracy of past forecasts at different geographical scales and over different time horizons can be visually assessed by varying the date from which to predict (to a date in the past) and comparing past predictions to the observed data.

#### Local predictors of transmission and importations

Spatial heterogeneity in case incidence, transmission and importation risk at the latest date of the forecasts is displayed in the app via three different maps showing the spatial distribution of cases at a NUTS-3 or NUTS-1/NUTS-2 level, the local risk of transmission (the population-weighted average value of the epidemic component for each region across all age groups), and the local risk of importation (the epidemic component for each region summed across age groups) (Figure 6). The local risk of transmission and importation are shown as a percentage of the highest value of the predictor in the country, and give insight into the spatial heterogeneity in risk in the country. In all three countries, the risk of transmission is more homogeneous across the regions than the risk of importation. The risk of importation, quantified by the endemic predictor, is heavily influenced by the number of inhabitants in a region, so the regions gathering most of the importation risks are regions containing the major cities in all three countries.

**Figure 6:**
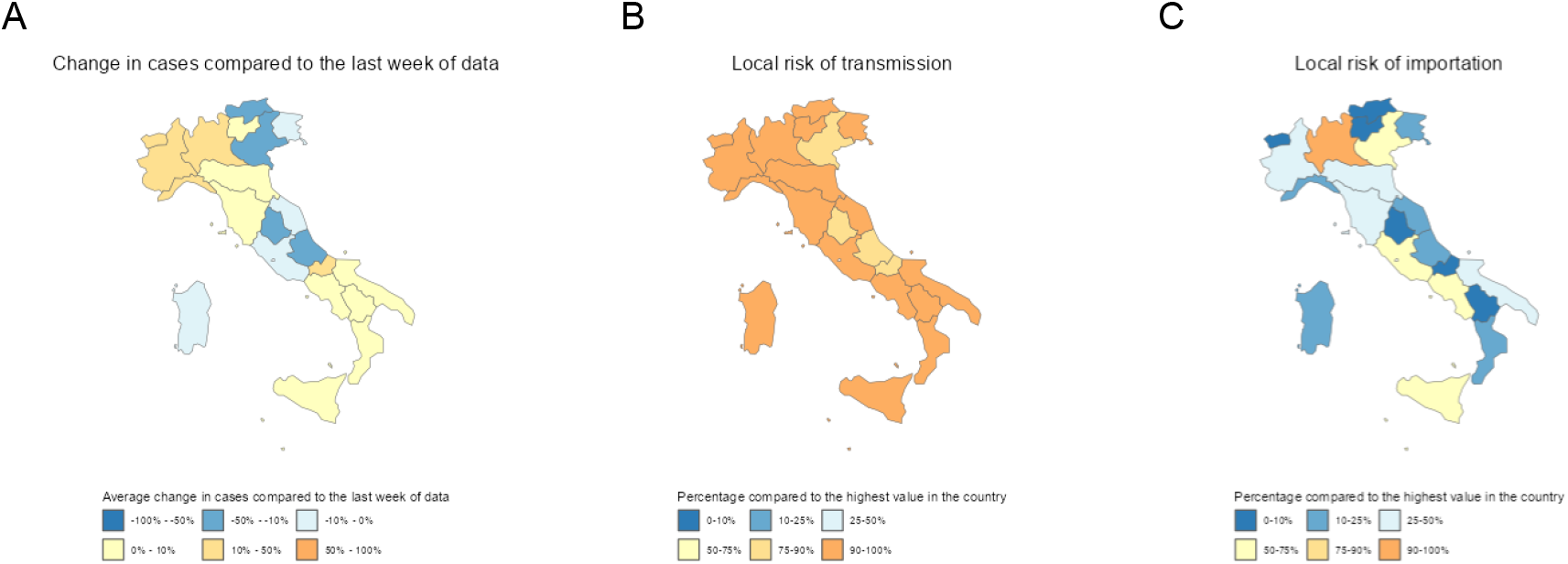
Visualisation of spatial heterogeneity in case incidence and risks of transmission and importations in Italy in the RShiny app. (A) Map of forecasted percentage change in cases in next week compared to the last week of data in Italy. (B) Map of local risk of transmission (as quantified by the estimated epidemic predictor in the model). (C) Map of local risk of importation (as quantified by the estimated endemic predictor in the model). All maps are at NUTS-2 level and show values on 7th March 2023.

#### Short-term transmission scenarios

Finally, simulations showing the impact of various changes in transmission (either increases due to changes in behaviour or new variants, or decreases caused by targeted or global NPIs) on predicted numbers of cases and deaths are shown in the app via similar figures to those used to display the case and death forecasts (Figure 7). Options for exploring the impact of changes in transmission include combinations of:

**Figure 7:**
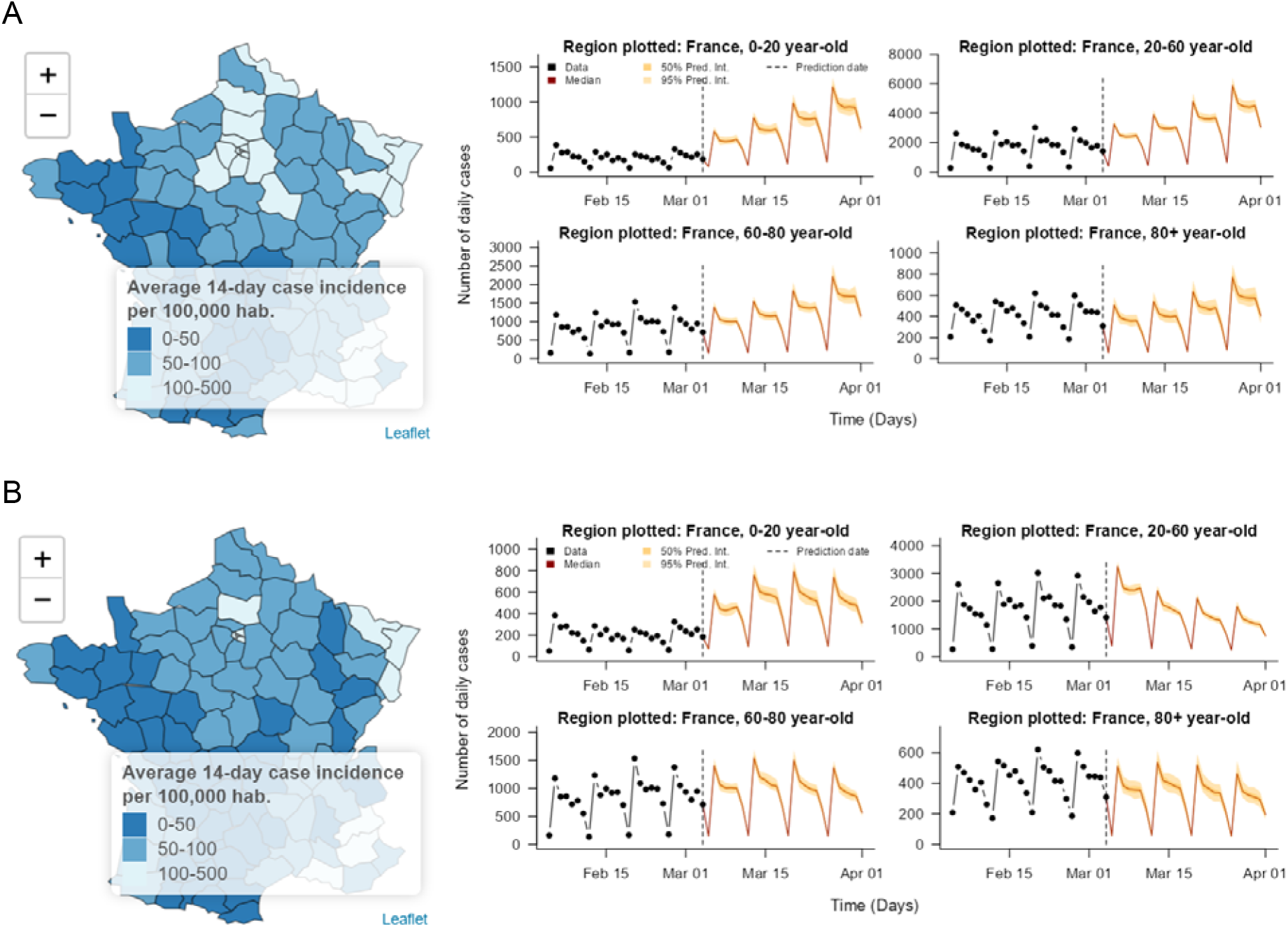
Visualisation of impact of different scenarios for changes in transmission (due to changes in behaviour, a new variant, or a change in NPIs) on forecasted cases in France in the RShiny app. Map of forecasted 14-day case incidence from 7th to 21st March 2023 at NUTS-3 level and age-stratified time series plots of national four-week-ahead case forecasts for (A) no change in transmission, and (B) a 40% decrease in transmission among 20-60-year-olds due to NPIs targeted at 20-60 year-olds with a one-week delay to effect.

- Increasing transmission by 0, 20, or 40% to represent different properties of an emerging variant, or changes in behaviour that lead to increased risks of spread.
- Dropping transmission by 0, 20, or 40% to represent the impact of increased NPIs.
- Targeting NPIs at the whole population or a specific age group.
- Removing importations from outside the selected country to represent stringent border closures.
- Changing the delay in the time it takes for NPIs to become effective (1 week or 2 weeks).

Throughout all three countries, removing importations has very little impact on the transmission dynamics in the case forecasts. Local transmission is sufficient to maintain transmission without new cases being added through the endemic component. On the other hand, the moderate and large changes in transmission through NPIs or changes in behaviour have a large impact on the forecasted number of cases and deaths. Delaying the implementation of NPIs by a week strongly decreases the impact of the control measures.

## Discussion

We have developed a framework to forecast subnational COVID-19 case and death incidence up to 4 weeks ahead, and explore the potential impact of changes in transmission on reported incidence. The framework was applied to France, Czechia and Italy. The model outputs are based on routinely collected, publicly available surveillance data. We have also developed a RShiny app, where users can visualise the 4-week-ahead forecasts of both reported cases and deaths, and the predicted impact of changes in transmission. The code we developed to implement the model and the RShiny App is publicly available in two Github repositories: https://github.com/EU-ECDC/RShinyCovidApp and https://github.com/EU-ECDC/BackendCovidApp. The model fits and scenario simulations are updated automatically every week.

Case and death forecasts aggregated at a country level perform comparably with the European COVID-19 Forecast Hub ensemble model, a benchmark in epidemic forecasting as an ensemble of forecasts from many independent models. In addition, our framework provides NUTS-3-region-level (i.e. much more highly spatially resolved) and, if age-stratified data is available, age-stratified forecasts. Therefore, it can be used for more targeted policy making and planning at a local level. Given the significant spatiotemporal heterogeneity in COVID-19 incidence, and changes in age patterns, the detailed local-level visualisation provided by our model is particularly useful to evaluate targeted control measures. This study highlights the need for surveillance systems that gather accurate, timely, age-stratified data, and the value of making such data publicly available to improve understanding and prediction of local transmission and outbreak response planning.

As demonstrated by our calibration analysis, the subnational and age-stratified case and death forecasts are accurate up to 2 weeks ahead: the case PIT histograms are flat for Italy and for most age groups for Czechia and France, while the median country-level forecast is closer to the data than the European COVID-19 Forecast Hub ensemble forecast. However, the forecasts become less reliable beyond a 2-week horizon. This may be due to fundamental predictability limits, the difficulty of forecasting changes in behaviour and/or sudden changes in transmission, e.g. due to the emergence of a new highly transmissible variant, more than a couple of weeks into the future or other factors that cause the model to be misspecified, and reflects similar findings from other forecasting efforts (7,17,57). If COVID-19 transmission dynamics become similar to those of seasonal influenza, sudden unexpected changes in transmission may become rarer, which would improve the performance of the model. The age-stratified PIT histograms in both Czechia and France show that calibration is especially difficult in younger age groups, where changes in case-finding strategy had a big impact that could not be fully captured by the testing covariates (58,59).

The model was built to provide accurate predictions of case and death incidence at a regional level. Our results show that generating reliable case forecasts several weeks ahead is challenging, even when using age-stratified local data. However, the accuracy of death forecasts was consistent throughout all the forecast horizons that were tested. The quality of the case and death forecasts was robust to the large reporting changes observed throughout the fitting period, and improved at the latest calibration dates where incidence was low in all three countries. This may be because local outbreaks in groups at risk in low-incidence settings are easier to forecast, rather than outbreaks where all regions are equally vulnerable. Forecasts past two weeks generated by the model appear to be overconfident, and underestimate the uncertainty in the potential level of transmissions. The alternative transmission scenarios in the Shiny App therefore help to illustrate the full range of variability in short-term regional transmission dynamics that is possible. The local risk of outbreaks and importations is also represented in the Shiny App, using the local predictors of the Endemic-Epidemic model. The forecasts, scenarios, and risk map constitute a reliable, thorough representation of the local risk of transmission.

The model fits and simulations highlight several features of COVID-19 dynamics common to all three countries: the risk of background importation of cases (i.e. new cases that were not linked to recent local infections) is always very strongly associated with the number of inhabitants in the region. Hence, more populated and urban areas are at higher risk of further importations of SARS-CoV-2 infections.. On the other hand, the risk of transmission is relatively homogeneous across regions, with no regions where the number of secondary cases expected is less than half the region most at risk (Figure 6B). This could change in future COVID-19 dynamics, where the local level of immunity may be sufficient to completely avoid transmission in certain regions, as is currently observed for pathogens such as measles (34). The vast majority of cases stem from the epidemic component of the Endemic-Epidemic model, showing that the local transmissibility is sufficient to maintain transmission, even without input from background importations.

Therefore, the removal of importations (e.g., by border closures) consistently resulted in a minimal impact on the expected number of case numbers up to four weeks ahead. In contrast, altering the transmission risk within the regions (e.g. via interventions targeted at specific age groups or encompassing the entire population) can substantially change the forecasted case dynamics. Even with strategies targeting a particular age group, we observe indirect effects on incidence in all age groups. We did not observe a strong effect of changes in transmission risk on the four-week-ahead death forecasts. However, we emphasise that we considered a three-week delay between cases and deaths and, hence, the impact of changes in transmission on deaths would be visible for longer-term death forecasts (beyond four weeks ahead).

### Including other countries

The framework is currently implemented for France, Czechia and Italy, but other countries can be straightforwardly incorporated, provided the data required for making the subnational daily case and weekly death forecasts (namely NUTS-3-level daily case counts, subnational daily death counts, NUTS-3-/NUTS-2-level daily/weekly vaccinations administered by dose, and NUTS-3-/NUTS-2-/national-level daily/weekly numbers tested) for those countries is available and up-to-date. Full instructions for incorporating a new country are available in the public GitHub repository for the model code: https://github.com/EU-ECDC/BackendCovidApp.

### Age stratification

Our comparison of forecasted and observed numbers of cases and deaths over all age groups for France and Czechia indicates that the non-age-stratified model has similar predictive performance in terms of overall cases/deaths as the age-stratified model (Supplementary Figures 6-9), albeit with slightly worse prediction of deaths (Supplementary Figure 9). We believe the loss in performance observed in death forecasts to be due to changes in the age structure of reported cases from changes in case detection. Indeed, active case finding may lead to milder cases being reported, while interruption of such a strategy would mean that a larger proportion of the cases are severe. Such changes are likely to be reflected in the age structure of the cases and such information is lost in the non-age-stratified version of the model.

The overall good performance of the non-age-stratified model is encouraging for the application of the model to other countries, very few of which report age-stratified case and death data. While we did not observe a significant improvement by age-stratifying the model, we emphasise that age-stratified data and models are crucial for evaluating the impact of targeted (age-specific) non-pharmaceutical and pharmaceutical interventions, as well as for health-economic analyses (e.g., the computation of DALYs).

### Limitations

Our forecasting framework does have some limitations. First, as for all forecasting studies, the forecasts are only as reliable as the input data, and we cannot guarantee the accuracy of the data imported from public data sources, which may have reporting errors or biases that we are not able to account for. The framework will only work as long as subnational (and age-stratified, for France and Czechia) case and death data continues to be reported online in the same location and format as it is currently, but several countries have ceased reporting subnational case and death data or changed the format or location of their reporting since we started developing the framework. Changes in data availability could potentially be addressed via changing spatial resolution of the model, but this would involve substantial modification of the model structure. We only forecast reported cases, which reflect a combination of underlying incidence of infections and reporting, rather than the “true” number of cases, but these still provide a useful indication of changes in transmission. The model for the death forecasts, which uses an estimate of the recent CFR (with uncertainty) to predict deaths, is relatively simple and assumes a constant relationship between changes in case numbers and changes in the CFR over time. However, the calibration analysis shows it provides a relatively straightforward and reliable means of translating case forecasts into death forecasts.

The contact matrices used to fit the model for France and Czechia were taken from pre-pandemic studies, and may not reflect the contact patterns between age groups in 2020 and 2021. Ideally, time-varying matrices would have been used to estimate age-stratified contact, but such matrices were not available or straightforward to implement in the Epidemic-Endemic framework (60). The addition of an age-specific intercept in the epidemic component allowed the model to modify the risk of transmission in each group where the number of contacts was not in line with the case counts. However, since these coefficients were not time-dependent, the model considers this age-stratified risk of new cases to be constant (if all other covariates do not vary).

We do not include information on NPIs or population mobility in the covariates in the model despite the improvements in predictions these may yield, since centralised databases of these covariates (such as Google Mobility data (61), the Oxford Stringency Index (62), and the ECDC-JRC Response Measures Database (2)) are no longer being updated, and binary covariates made the model less consistent and comparable between countries, while potentially not being specific enough (i.e. they incorporated the impact of other parameters associated with transmission). Including more NPI covariates also led to issues with identifying the effects of different interventions as many were implemented at the same or overlapping times.

### Generalising beyond COVID-19

The flexible framework developed in this paper could be used or readily adapted to model incidence of both novel and seasonal pathogens of public health importance, such as influenza, in order to predict local health burden and inform outbreak response. The Endemic-Epidemic framework underlying our model has already been applied to a variety of other pathogens including measles, cholera, leishmaniasis and pertussis (34,63–65). However, the number of model parameters that can be estimated directly depends on the amount of data available.

Therefore, in its current specification (with a large number of parameters), the model may not be suitable for the early stages of an outbreak (except if the pathogen is seasonal, with available data on previous outbreaks). However, it can be used to estimate the impact of various covariates (from different data sources) on transmission risk.

## Data availability

All source data used in the study were openly available. The code used to develop the analysis and links to the data sources are available on Github (back-end code: https://github.com/EU-ECDC/BackendCovidApp, code for the RShiny App: https://github.com/EU-ECDC/RShinyCovidApp).

## Author’s contribution

AR, LACC, AJK and SF led on conceptualisation and methodology. AR and LACC were in charge of software, validation, formal analysis, data curation, visualisation, writing - original draft preparation, and writing - review and editing. BP, RN, FS, and RG were in charge of project administration, funding acquisition, and writing - review and editing. AJK and SF were responsible for supervision and writing - review and editing.

## Acknowledgments

We acknowledge Dr Naomi Waterlow (London School of Hygiene and Tropical Medicine) and Dr Sophie Meakin (Epicentre, Paris) for their feedback on early versions of the RShiny App, which helped us clarify the presentation of the results. We also acknowledge Dr Johannes Bracher (Karlsruhe Institute of Technology) for helpful discussions regarding the hhh4addon and hhh4contacts R packages.

## References

1. Chapman LAC, Barnard RC, Russell TW, Abbott S, van Zandvoort K, Davies NG, et al. Unexposed populations and potential COVID-19 hospitalisations and deaths in European countries as per data up to 21 November 2021. Eurosurveillance. 2022 Jan 6;27(1):2101038.

2. Response Measures Database (RMD) [Internet]. European Centre for Disease Prevention and Control. 2022 [cited 2023 May 19]. Available from: https://www.ecdc.europa.eu/en/publications-data/response-measures-database-rmd

3. Davies NG, Abbott S, Barnard RC, Jarvis CI, Kucharski AJ, Munday JD, et al. Estimated transmissibility and impact of SARS-CoV-2 lineage B.1.1.7 in England. Science [Internet]. 2021 Apr 9;372(6538). Available from: http://dx.doi.org/10.1126/science.abg3055

4. Sonabend R, Whittles LK, Imai N, Perez-Guzman PN, Knock ES, Rawson T, et al. Non-pharmaceutical interventions, vaccination, and the SARS-CoV-2 delta variant in England: a mathematical modelling study. Lancet. 2021 Nov 13;398(10313):1825–35.

5. Birrell P, Blake J, van Leeuwen E, Gent N, De Angelis D. Real-time nowcasting and forecasting of COVID-19 dynamics in England: the first wave. Philos Trans R Soc Lond B Biol Sci. 2021 Jul 19;376(1829):20200279.

6. Friedman J, Liu P, Troeger CE, Carter A, Reiner RC, Barber RM, et al. Predictive performance of international COVID-19 mortality forecasting models. Nat Commun. 2021 May 10;12(1):1–13.

7. Funk S, Abbott S, Atkins BD, Baguelin M, Baillie JK, Birrell P, et al. Short-term forecasts to inform the response to the Covid-19 epidemic in the UK [Internet]. medRxiv. 2020 [cited 2023 May 19]. p. 2020.11.11.20220962. Available from: https://www.medrxiv.org/content/10.1101/2020.11.11.20220962v2.abstract

8. Fonseca-Rodríguez O, Gustafsson PE, San Sebastián M, Connolly A-MF. Spatial clustering and contextual factors associated with hospitalisation and deaths due to COVID-19 in Sweden: a geospatial nationwide ecological study. BMJ Glob Health [Internet]. 2021 Jul;6(7). Available from: http://dx.doi.org/10.1136/bmjgh-2021-006247

9. Sartorius B, Lawson AB, Pullan RL. Modelling and predicting the spatio-temporal spread of COVID-19, associated deaths and impact of key risk factors in England. Sci Rep. 2021 Mar 8;11(1):5378.

10. Wang Q, Dong W, Yang K, Ren Z, Huang D, Zhang P, et al. Temporal and spatial analysis of COVID-19 transmission in China and its influencing factors. Int J Infect Dis. 2021 Apr;105:675–85.

11. Chinazzi M, Davis JT, Ajelli M, Gioannini C, Litvinova M, Merler S, et al. The effect of travel restrictions on the spread of the 2019 novel coronavirus (COVID-19) outbreak. Science. 2020 Apr 24;368(6489):395–400.

12. Danon L, Brooks-Pollock E, Bailey M, Keeling M. A spatial model of COVID-19 transmission in England and Wales: early spread, peak timing and the impact of seasonality. Philos Trans R Soc Lond B Biol Sci. 2021 Jul 19;376(1829):20200272.

13. Gatto M, Bertuzzo E, Mari L, Miccoli S, Carraro L, Casagrandi R, et al. Spread and dynamics of the COVID-19 epidemic in Italy: Effects of emergency containment measures. Proc Natl Acad Sci U S A. 2020 May 12;117(19):10484–91.

14. Pei S, Kandula S, Shaman J. Differential effects of intervention timing on COVID-19 spread in the United States. Sci Adv [Internet]. 2020 Dec;6(49). Available from: http://dx.doi.org/10.1126/sciadv.abd6370

15. Read JM, Bridgen JRE, Cummings DAT, Ho A, Jewell CP. Novel coronavirus 2019-nCoV (COVID-19): early estimation of epidemiological parameters and epidemic size estimates. Philos Trans R Soc Lond B Biol Sci. 2021 Jul 19;376(1829):20200265.

16. covid19uk [Internet]. GitLab. [cited 2023 May 19]. Available from: https://fhm-chicas-code.lancs.ac.uk/jewell/covid19uk

17. Cramer EY, Ray EL, Lopez VK, Bracher J, Brennen A, Castro Rivadeneira AJ, et al. Evaluation of individual and ensemble probabilistic forecasts of COVID-19 mortality in the United States. Proc Natl Acad Sci U S A. 2022 Apr 12;119(15):e2113561119.

18. stsmodel [Internet]. GitLab. [cited 2023 May 19]. Available from: https://gitlab.com/chicas-covid19/stsmodel

19. Giuliani D, Dickson MM, Espa G, Santi F. Modelling and predicting the spatio-temporal spread of cOVID-19 in Italy. BMC Infect Dis. 2020 Sep 23;20(1):700.

20. Douwes-Schultz D, Sun S, Schmidt AM, Moodie EEM. Extended Bayesian endemic-epidemic models to incorporate mobility data into COVID-19 forecasting. Can J Stat. 2022 Sep;50(3):713–33.

21. Fronterre C, Read JM, Rowlingson B, Bridgen J, Alderton S, Diggle PJ, et al. COVID-19 in England: spatial patterns and regional outbreaks [Internet]. bioRxiv. medRxiv; 2020. Available from: http://medrxiv.org/lookup/doi/10.1101/2020.05.15.20102715

22. Ssentongo P, Fronterre C, Geronimo A, Greybush SJ, Mbabazi PK, Muvawala J, et al. Pan-African evolution of within- and between-country COVID-19 dynamics. Proc Natl Acad Sci U S A [Internet]. 2021 Jul 13;118(28). Available from: http://dx.doi.org/10.1073/pnas.2026664118

23. Dickson MM, Espa G, Giuliani D, Santi F, Savadori L. Assessing the effect of containment measures on the spatio-temporal dynamic of COVID-19 in Italy. Nonlinear Dyn. 2020 Aug 8;101(3):1833–46.

24. Grimée M, Bekker-Nielsen Dunbar M, Hofmann F, Held L, SUSPend modelling consortium. Modelling the effect of a border closure between Switzerland and Italy on the spatiotemporal spread of COVID-19 in Switzerland. Spat Stat. 2022 Jun;49:100552.

25. Berlemann M, Haustein E. Right and Yet Wrong: A Spatio-Temporal Evaluation of Germany’s COVID-19 Containment Policy [Internet]. 2020 [cited 2023 May 22]. Available from: https://papers.ssrn.com/abstract=3662054

26. Fritz C, Kauermann G. On the interplay of regional mobility, social connectedness and the spread of COVID-19 in Germany. J R Stat Soc Ser A Stat Soc. 2022 Jan;185(1):400–24.

27. Bekker-Nielsen Dunbar M, Held L. Endemic-Epidemic Framework Used in Covid-19 Modelling. Revstat Stat J. 2020 Oct 20;18(5):565–74.

28. Celani A, Giudici P. Endemic-epidemic models to understand COVID-19 spatio-temporal evolution. Spat Stat. 2022 Jun;49:100528.

29. Meyer S, Held L, Höhle M. Spatio-temporal analysis of epidemic phenomena using the R package surveillance. J Stat Softw [Internet]. 2017;77(11). Available from: http://www.jstatsoft.org/v77/i11/

30. Temporal and Spatio-Temporal Modeling and Monitoring of Epidemic Phenomena [R package surveillance version 1.21.1]. 2023 May 19 [cited 2023 May 19]; Available from: https://CRAN.R-project.org/package=surveillance

31. Bracher J, Ray EL, Gneiting T, Reich NG. Evaluating epidemic forecasts in an interval format. PLoS Comput Biol. 2021 Feb;17(2):e1008618.

32. Bracher J, Held L. hhh4addon: extending the functionality of surveillance: hhh4. R package. 2019.

33. Bracher J, Held L. Endemic-epidemic models with discrete-time serial interval distributions for infectious disease prediction. Int J Forecast. 2022 Jul;38(3):1221–33.

34. Robert A, Kucharski AJ, Funk S. The impact of local vaccine coverage and recent incidence on measles transmission in France between 2009 and 2018. BMC Med. 2022 Mar 10;20(1):77.

35. Alene M, Yismaw L, Assemie MA, Ketema DB, Gietaneh W, Birhan TY. Serial interval and incubation period of COVID-19: a systematic review and meta-analysis. BMC Infect Dis. 2021 Mar 11;21(1):257.

36. Pung R, Mak TM, CMMID COVID-19 working group, Kucharski AJ, Lee VJ. Serial intervals in SARS-CoV-2 B.1.617.2 variant cases. Lancet. 2021 Sep 4;398(10303):837–8.

37. Nishiura H, Linton NM, Akhmetzhanov AR. Serial interval of novel coronavirus (COVID-19) infections [Internet]. Vol. 93, International Journal of Infectious Diseases. 2020. p. 284–6. Available from: http://dx.doi.org/10.1016/j.ijid.2020.02.060

38. Meyer S. Age-Structured Spatio-Temporal Models for Infectious Disease Counts [R package hhh4contacts version 0.13.1]. 2020 Mar 20 [cited 2023 May 19]; Available from: https://CRAN.R-project.org/package=hhh4contacts

39. Meyer S, Held L. Incorporating social contact data in spatio-temporal models for infectious disease spread. Biostatistics. 2017 Apr 1;18(2):338–51.

40. Béraud G, Kazmercziak S, Beutels P, Levy-Bruhl D, Lenne X, Mielcarek N, et al. The French Connection: The First Large Population-Based Contact Survey in France Relevant for the Spread of Infectious Diseases. PLoS One. 2015 Jul 15;10(7):e0133203.

41. Prem K, van Zandvoort K, Klepac P, Eggo RM, Davies NG, Centre for the Mathematical Modelling of Infectious Diseases COVID-19 Working Group, et al. Projecting contact matrices in 177 geographical regions: An update and comparison with empirical data for the COVID-19 era. PLoS Comput Biol. 2021 Jul;17(7):e1009098.

42. Linton NM, Kobayashi T, Yang Y, Hayashi K, Akhmetzhanov AR, Jung S-M, et al. Incubation Period and Other Epidemiological Characteristics of 2019 Novel Coronavirus Infections with Right Truncation: A Statistical Analysis of Publicly Available Case Data. J Clin Med Res [Internet]. 2020 Feb 17;9(2). Available from: http://dx.doi.org/10.3390/jcm9020538

43. Khalili M, Karamouzian M, Nasiri N, Javadi S, Mirzazadeh A, Sharifi H. Epidemiological characteristics of COVID-19: a systematic review and meta-analysis. Epidemiol Infect. 2020 Jun 29;148:e130.

44. Challen R, Brooks-Pollock E, Tsaneva-Atanasova K, Danon L. Meta-analysis of the severe acute respiratory syndrome coronavirus 2 serial intervals and the impact of parameter uncertainty on the coronavirus disease 2019 reproduction number. Stat Methods Med Res. 2022 Sep;31(9):1686–703.

45. European Centre for Disease Prevention and Control. Data on SARS-CoV-2 variants in the EU/EEA [Internet]. [cited 2022 Jul 26]. Available from: https://www.ecdc.europa.eu/en/publications-data/data-virus-variants-covid-19-eueea

46. Eurostat. Methodology - Rural development [Internet]. [cited 2022 Jul 26]. Available from: https://ec.europa.eu/eurostat/web/rural-development/methodology

47. World Health Organization. Daily cases and deaths by date reported to WHO [Internet]. 2023 [cited 2023 Jan 30]. Available from: https://covid19.who.int/WHO-COVID-19-global-data.csv

48. Santé publique France. Données de laboratoires pour le dépistage [Internet]. [cited 2022 Jul 26]. Available from: https://www.data.gouv.fr/fr/datasets/donnees-de-laboratoires-pour-le-depistage-a-compter-du-18-05-2022-si-dep/

49. Ministerstvo Zdravotnictví České republiky. COVID-19 in the Czech Republic: Open data sets and downloadable sets [Internet]. [cited 2022 Jul 26]. Available from: https://onemocneni-aktualne.mzcr.cz/api/v2/covid-19

50. Dipartimento della Protezione Civile. Dati COVID-19 Italia [Internet]. [cited 2022 Jul 26]. Available from: https://github.com/pcm-dpc/COVID-19

51. datavaccin-covid. Données vaccination par tranche d’âge, type de vaccin et département / région [Internet]. [cited 2022 Jul 26]. Available from: https://datavaccin-covid.ameli.fr/explore/dataset/donnees-vaccination-par-tranche-dage-type-de-vaccin-et-departement/information/

52. European Centre for Disease Prevention and Control. Data on COVID-19 vaccination in the EU/EEA [Internet]. [cited 2022 Jul 26]. Available from: https://www.ecdc.europa.eu/en/publications-data/data-covid-19-vaccination-eu-eea

53. Insee. Estimation de la population au 1er janvier 2022 [Internet]. [cited 2022 Jul 26]. Available from: https://www.insee.fr/fr/statistiques/1893198

54. Eurostat. Population on 1 January by age group, sex and NUTS 3 region [Internet]. [cited 2023 Jan 2]. Available from: https://ec.europa.eu/eurostat/web/products-datasets/product?code=demo_r_pjangrp3

55. O. Wahltinez and others. COVID-19 Open-Data: curating a fine-grained, global-scale data repository for SARS-CoV-2. 2020; Available from: https://goo.gle/covid-19-open-data

56. Czado C, Gneiting T, Held L. Predictive model assessment for count data. Biometrics. 2009 Dec;65(4):1254–61.

57. Sherratt K, Gruson H, Grah R, Johnson H, Niehus R, Prasse B, et al. Predictive performance of multi-model ensemble forecasts of COVID-19 across European nations [Internet]. 2022. Available from: http://medrxiv.org/lookup/doi/10.1101/2022.06.16.22276024

58. Leng T, Hill EM, Holmes A, Southall E, Thompson RN, Tildesley MJ, et al. Quantifying pupil-to-pupil SARS-CoV-2 transmission and the impact of lateral flow testing in English secondary schools. Nat Commun. 2022 Mar 1;13(1):1106.

59. Paltiel AD, Zheng A, Walensky RP. Assessment of SARS-CoV-2 Screening Strategies to Permit the Safe Reopening of College Campuses in the United States. JAMA Netw Open. 2020 Jul 1;3(7):e2016818.

60. Munday JD, Abbott S, Meakin S, Funk S. Evaluating the use of social contact data to produce age-specific forecasts of SARS-CoV-2 incidence [Internet]. bioRxiv. 2022. Available from: https://www.medrxiv.org/content/10.1101/2022.12.02.22282935.abstract

61. Google. COVID-19 Community Mobility Reports [Internet]. [cited 2023 May 19]. Available from: https://www.google.com/covid19/mobility/

62. Hale T, Angrist N, Goldszmidt R, Kira B, Petherick A, Phillips T, et al. A global panel database of pandemic policies (Oxford COVID-19 Government Response Tracker). Nat Hum Behav. 2021 Apr;5(4):529–38.

63. Nightingale ES, Chapman LAC, Srikantiah S, Subramanian S, Jambulingam P, Bracher J, et al. A spatio-temporal approach to short-term prediction of visceral leishmaniasis diagnoses in India. PLoS Negl Trop Dis. 2020 Jul;14(7):e0008422.

64. Munro AD, Smallman-Raynor M, Algar AC. Long-term changes in endemic threshold populations for pertussis in England and Wales: A spatiotemporal analysis of Lancashire and South Wales, 1940-69. Soc Sci Med. 2021 Nov;288:113295.

65. Jeandron A. Tap water access and its relationship with cholera and other diarrhoeal diseases in an urban, cholera-endemic setting in the Democratic Republic of the Congo [Internet] [doctoral]. London School of Hygiene & Tropical Medicine; 2020 [cited 2023 May 22]. Available from: https://researchonline.lshtm.ac.uk/id/eprint/4659288/

