## Supplementary Material for "Predicting subnational incidence of COVID-19 cases and deaths in EU countries"

### Distributed lags using SARS-CoV-2 serial interval

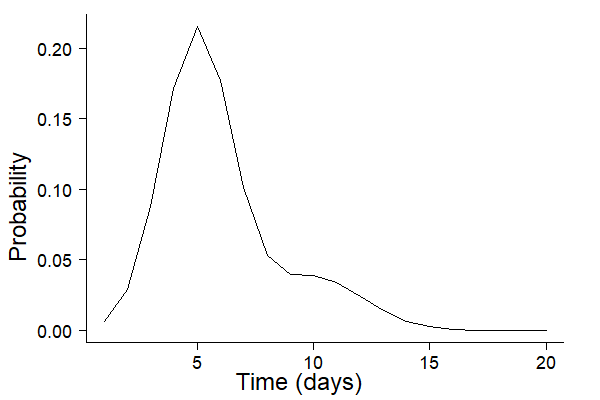

Supplementary Figure 1: Lag distribution used in the epidemic component in the model. The lag distribution determines the weighting of the effect of cases on previous days on cases on the current day.

### Model equation for each country

#### France

We implemented an age-stratified Endemic-Epidemic model based on Equation (A1):

$log(\phi_{ait})=\alpha^{(\phi)}+\alpha_{a}^{(\phi)}+\beta_{pop\_age}^{(\phi)}log(po{p\_age}_{ai}) + \beta_{pop\_tot}^{(\phi)}log(pop_{i})$	(A1)

$+\beta_{tue}tue_{t}+\beta_{wed}wed+\beta_{thu}thu_{t}+\beta_{fri}fri_{t}+\beta_{sat}sat_{t}+\beta_{sun}sun_{t}$

$+\beta_{test\_prop}log(tes{t\_prop}_{t})+\beta_{test\_age}log(tes{t\_age}_{ait})$

$+\beta_{rural}^{(\phi)}rural_{i}+\beta_{int\_rur}^{(\phi)}int\_rur_{i}+\beta_{int\_urb}^{(\phi)}int\_ urb_{i}$

$+\beta_{cov}^{(\phi)}log(1-cov_{ait})+\beta_{inc\_old}^{(\phi)}log(1-in{c\_old}_{ait})+\beta_{inc\_new}^{(\phi)}log(1-in{c\_new}_{ait})$

$+\beta_{delta}delta_{t}+\beta_{omicron}omicron_{t}$

$+\beta_{sin}^{(\phi)}sin(2*\pi*t/365)+\beta_{cos}^{(\phi)}cos(2*\pi*t/365)$

$log(\nu_{ait})={\alpha^{(\nu)}+}\beta_{pop}^{(\nu)}log(pop_{i} * po{p\_age}_{ai})$	$+\beta_{rural}^{(\nu)}rural_{i}+\beta_{int\_rur}^{(\nu)}int\_rur_{i}+\beta_{int\_urb}^{(\nu)}int\_urb_{i}$

$+\beta_{cases\_eur}log(cases\_eur_{t})$

$+\beta_{sin}^{(\nu)}sin(2*\pi*t/365)+\beta_{cos}^{(\nu)}cos(2*\pi*t/365)$.

Here, $\alpha^{(\phi)}$ and $\alpha^{(\nu)}$ are fixed intercepts for the epidemic and endemic components, with age adjustments $\alpha_{a}^{(\phi)}$ in the epidemic component. All covariates are defined in Supplementary Table 1. The cumulative incidence of cases by age and region between 1 month before day $t$ and the start of the fitting period, $inc\_old_{ait}$, and in the last month, $inc\_new_{ait}$, were introduced to allow the model to better capture short- and medium-term changes in immunity due to differences in the rate of spread of different variants (e.g. Delta and Omicron). The contact matrix between age groups was taken from Béraud et al (1).

#### Czechia

$log(\phi_{ait})=\alpha^{(\phi)}+\alpha_{a}^{(\phi)}+\beta_{pop\_age}^{(\phi)}log(po{p\_age}_{ai}) + \beta_{pop\_tot}^{(\phi)}log(pop_{i})$	(A2)

$+\beta_{tue}tue_{t}+\beta_{wed}wed+\beta_{thu}thu_{t}+\beta_{fri}fri_{t}+\beta_{sat}sat_{t}+\beta_{sun}sun_{t}$

$+\beta_{test\_prop}log(tes{t\_prop}_{t})+\beta_{test\_age}log(tes{t\_age}_{at})$

$+\beta_{int\_rur}^{(\phi)}int\_rur_{i}+\beta_{int\_urb}^{(\phi)}int\_ urb_{i}$

$+\beta_{cov}^{(\phi)}log(1-cov_{ait})+\beta_{inc\_old}^{(\phi)}log(1-in{c\_old}_{ait})+\beta_{inc\_new}^{(\phi)}log(1-in{c\_new}_{ait})$

$+\beta_{delta}delta_{t}+\beta_{omicron}omicron_{t}$

$+\beta_{sin}^{(\phi)}sin(2*\pi*t/365)+\beta_{cos}^{(\phi)}cos(2*\pi*t/365)$

$log(\nu_{ait})={\alpha^{(\nu)}+}\beta_{pop}^{(\nu)}log(pop_{i} * pop_{ai})$ $+\beta_{int\_rur}^{(\nu)}int\_rur_{i}+\beta_{int\_urb}^{(\nu)}int\_urb_{i}$

$+\beta_{cases\_eur}log(cases\_eur_{t})$

$+\beta_{sin}^{(\nu)}sin(2*\pi*t/365)+\beta_{cos}^{(\nu)}cos(2*\pi*t/365)$.

In comparison to France, since there was no NUTS-3 region in Czechia classified as “rural”, this level was dropped from the equation. The contact matrix was extracted from Prem et al (2).

#### Italy

$log(\phi_{ait})=\alpha^{(\phi)}+ \beta_{pop\_tot}^{(\phi)}log(pop_{i})$	(A3)

$+\beta_{tue}tue_{t}+\beta_{wed}wed+\beta_{thu}thu_{t}+\beta_{fri}fri_{t}+\beta_{sat}sat_{t}+\beta_{sun}sun_{t}$

$+\beta_{test\_prop}log(test\_prop_{t})$

$+\beta_{rural}^{(\phi)}rural_{i}+\beta_{int\_rur}^{(\phi)}int\_ rur_{i}+\beta_{int\_urb}^{(\phi)}int\_ urb_{i}$

$+\beta_{cov}^{(\phi)}log(1-cov_{it})+\beta_{inc\_old}^{(\phi)}log(1-in{c\_old}_{it})+\beta_{inc\_new}^{(\phi)}log(1-in{c\_new}_{it})$

$+\beta_{delta}delta_{t}+\beta_{omicron}omicron_{t}$

$+\beta_{sin}^{(\phi)}sin(2*\pi*t/365)+\beta_{cos}^{(\phi)}cos(2*\pi*t/365)$

$log(\nu_{ait})={\alpha^{(\nu)}+}\beta_{pop}^{(\nu)}log(pop_{i} )$	$+\beta_{rural}^{(\nu)}rural_{i}+\beta_{int\_rur}^{(\nu)}int\_rur_{i}+\beta_{int\_urb}^{(\nu)}int\_urb_{i}$

$+\beta_{cases\_eur}log(cases\_eur_{t})$

$+\beta_{sin}^{(\nu)}sin(2*\pi*t/365)+\beta_{cos}^{(\nu)}cos(2*\pi*t/365)$.

Since age-stratified data was not available in Italy, the model described in Equation (A3) does not include an age-specific intercept, nor the covariates $test\_age$ and $po{p\_age}_{ai}$ (Supplementary Table 1). The covariate $test\_prop$ now describes the proportion of the overall national population tested in the past 14 days.

**Supplementary Table 1: Definitions and sources of covariates included in the model**

| **Variable** | **Definition** | **Source** |
| --- | --- | --- |
| $pop_{i}$ | Total population of region $i$ (in 100,000s) | Country dependent, see Table 1 |
| $po{p\_age}_{ai}$ | Proportion of population of region $i$ that is in age group $a$ | Country dependent, see Table 1 |
| $tue_{t}$, $wed_{t}$, $thu_{t}$, $fri_{t}$,$sat_{t}$,$sun_{t}$ | Binary indicator variable controlling for the day of the week effect | n/a |
| $tes{t\_prop}_{t}$ | Proportion of the population tested in the last 14 days nationally | Country dependent, see Table 1 |
| $tes{t\_age}_{a(i)t}$ | Proportion of the population in age group $a$ (in region $i$ if subnational data is available) tested in the last 14 days | Country dependent, see Table 1 |
| $rural_{i}$ | Binary indicator variable for whether region $i$ is rural | (3) |
| $int\_rural_{i}$ | Binary indicator variable for whether region  $i$ is predominantly rural | (3) |
| $int\_urb_{i}$ | Binary indicator variable for whether region  $i$ is predominantly urban | (3) |
| $cov_{ait}$ | where $cov_{ait}$ is the proportion of the population of age group $a$ in region $i$ who have received three vaccine doses before day $t$, or have their second dose in the past 120 days. | Country dependent, see Table 1 |
| $in{c\_old}_{ait}$ | cumulative incidence of cases in age group $a$ in region $i$ from between the start of the fitting period and 1 month ago | Country dependent, see Table 1 |
| $in{c\_new}_{ait}$ | cumulative incidence of cases in age group $a$ in region $i$ in the last month (from day $t-30$ up to day $t-1$) | Country dependent, see Table 1 |
| $delta_{t}$ | Binary indicator variable for whether day $t$ was in a period when the proportion of sequenced cases that were Delta was higher than 30% | (4) |
| $omicron_{t}$ | Binary indicator variable for whether day $t$ was in a period when the proportion of sequenced cases that were Omicron was higher than 30% | (4) |
| $cases\_eur_{t}$ | Number of cases in the rest of Europe in the past 30 days on day $t$ (in 100,000s) | (4) |

### Model equation for non-age-stratified model for France and Czechia

For the non-age-stratified models for France and Czechia we drop the age-dependent covariates from the linear predictors for the epidemic and endemic components, such that the equations for France are:

$log(\phi_{it})=\alpha^{(\phi)}+ \beta_{pop\_tot}^{(\phi)}log(pop_{i})$	(A4)

$+\beta_{tue}tue_{t}+\beta_{wed}wed+\beta_{thu}thu_{t}+\beta_{fri}fri_{t}+\beta_{sat}sat_{t}+\beta_{sun}sun_{t}$

$+\beta_{test\_prop}log(tes{t\_prop}_{t})$

$+\beta_{rural}^{(\phi)}rural_{i}+\beta_{int\_rur}^{(\phi)}int\_rur_{i}+\beta_{int\_urb}^{(\phi)}int\_ urb_{i}$

$+\beta_{cov}^{(\phi)}log(1-cov_{it})+\beta_{inc\_old}^{(\phi)}log(1-in{c\_old}_{it})+\beta_{inc\_new}^{(\phi)}log(1-in{c\_new}_{it})$

$+\beta_{delta}delta_{t}+\beta_{omicron}omicron_{t}$

$+\beta_{sin}^{(\phi)}sin(2*\pi*t/365)+\beta_{cos}^{(\phi)}cos(2*\pi*t/365)$

$log(\nu_{it})={\alpha^{(\nu)}+}\beta_{pop}^{(\nu)}log(pop_{i})$	$+\beta_{rural}^{(\nu)}rural_{i}+\beta_{int\_rur}^{(\nu)}int\_rur_{i}+\beta_{int\_urb}^{(\nu)}int\_urb_{i}$

$+\beta_{cases\_eur}log(cases\_eur_{t})$

$+\beta_{sin}^{(\nu)}sin(2*\pi*t/365)+\beta_{cos}^{(\nu)}cos(2*\pi*t/365)$.

and those for Czechia are the same except without the $rural_{i}$ covariate as there are no regions classified as “rural” in Czechia.

#### Definition of predictive performance scores

##### Rank probability score (RPS)

The ranked probability score (RPS) for integer forecasts is defined as:

$RPS(P,y)=\sum_{k=0}^{\infty} {(F}_{P}(k)-1(y \leq k))^{2}$

where $y$ is the true value, $P$ is the predictive distribution, $F_{P}$ is its cumulative distribution function and $1()$ is the indicator function (5). It compares the cumulative distribution of $P$ to that of an ideal forecast with all probability mass assigned to the observed outcome $y$, and thus takes the entire predictive distribution into account. Smaller values of the RPS indicate better predictive performance.

##### Dawid-Sebastiani score (DSS)

The Dawid-Sebastiani score (DSS) is based on the first two moments of the predictive distribution of $P$ and is defined as:

$DSS(P,y)=\left( \frac{y-\mu_{P}}{\sigma_{P}} \right)^{2}+2log\sigma_{P}$

Smaller values of the DSS indicate better calibration.

##### Squared error score (SES)

The squared error score (SES) is defined as the squared error of the mean of the predictive distribution $\mu_{P}$:

$SES(P,y)=(y-\mu_{P})^{2}$

Smaller SES values indicate better predictive performance.

##### Weighted interval score (WIS)

The weighted interval score (WIS) for quantile forecasts is composed of three components: a sharpness component and separate penalties for over- and under-prediction. For a single interval, it is computed as:

$IS_{\alpha}(P,y)=(u-l)+\frac{2}{\alpha}1(y \leq l) +\frac{2}{\alpha}1(y \geq u)$

where $l$ and $u$ are the $\frac{\alpha}{2}$ and $1-\frac{\alpha}{2}$ quantiles of $P$. For a set of $K$ prediction intervals and the median $m$, it is calculated as a weighted sum:

$WIS_{\alpha}(P,y)=\frac{1}{K+1/2}(w_{0}|y-m|+\sum_{k=1}^{K} w_{k}IS_{\alpha_{k}}(P,y))$

with non-negative weights $w_{0},w_{1}, ...,w_{K}$ (6). We choose $w_{0}=\frac{1}{2}$ and $w_{k}=\frac{\alpha_{k}}{2}$, which means that the WIS converges to the rank probability score (RPS) as the number of intervals increases. Like the RPS, smaller scores indicate better predictive performance.

##### Squared error of the median

The squared error of the median is a scoring rule for point forecasts and is defined as:

$SE_{m}(P,y)=(y-m)^{2}$

### Calibration: Comparison with baseline model

Supplementary Table 2: Median and 95% interval of ranked probability score (RPS), Dawid-Sebastiani score (DSS) and squared error score (SES) of one-, two-, three- and four-week-ahead local age-stratified case forecasts for Czechia and France and non-age-stratified forecasts for Italy for the baseline Endemic-Epidemic model (without transmission between regions, covariates or seasonality) and full Endemic-Epidemic model (with covariates and seasonality) across all time points in the prediction period from 29 October 2022 to 22 April 2023

| Country | Forecast horizon (weeks ahead) | RPS, median (95% interval) | | DSS, median (95% interval) | | SES, median (95% interval) | |
| --- | --- | --- | --- | --- | --- | --- | --- |
|  |  | Baseline | Full | Baseline | Full | Baseline | Full |
| Czechia | 1 | 3.39 (1.56-7.41) | 1.57 (0.368-4.05) | 4.94 (3.17-8.56) | 2.98 (0.423-5.03) | 54.5 (8.6-275) | 9.3 (0.755-57.4) |
|  | 2 | 5.50 (1.58-12.3) | 1.77 (0.37-4.81) | 6.06 (3.33-10.2) | 3.77 (0.515-7.39) | 156 (9.61-869) | 12 (0.81-79.3) |
|  | 3 | 8.82 (1.67-22.7) | 2.01 (0.358-5.48) | 7.07 (3.38-9.37) | 5.65 (0.486-12.9) | 415 (9.34-2840) | 15.1 (0.806-99.1) |
|  | 4 | 14.2 (1.65-39.1) | 2.2 (0.357-6.21) | 7.92 (3.33-9.87) | 8.66 (0.578-24.5) | 1080 (9.43-8880) | 17.6 (0.685-123) |
| France | 1 | 7.12 (1.88-27.3) | 4.2 (1.23-18.4) | 5.47 (3.06-8.73) | 4.41 (1.87-7.88) | 293 (22.8-5040) | 103 (9.55-2250) |
|  | 2 | 9.09 (2.56-38.1) | 6.57 (1.76-33.8) | 6.07 (3.69-11.9) | 5.22 (2.32-11.9) | 460 (35.7-9320) | 264 (19.7-7510) |
|  | 3 | 11 (3.03-49.5) | 9.71 (2.54-54.6) | 6.57 (4.21-14.1) | 6.32 (2.97-17.2) | 735 (43.1-17500) | 559 (36.6-18100) |
|  | 4 | 13.6 (3.35-62.9) | 13.6 (3.44-80.5) | 7.02 (4.54-14.6) | 7.48 (3.4-27.2) | 1130 (57.4-29700) | 1010 (69.2-34400) |
| Italy | 1 | 25.6 (7.09-132) | 22 (6.5-122) | 8.02 (5.84-11.1) | 7.61 (5.49-10.7) | 3300 (271-104000) | 2390 (218-96300) |
|  | 2 | 28.7 (8.3-147) | 23.6 (7.18-135) | 8.43 (6.16-11.5) | 7.99 (5.74-11.8) | 4360 (367-133000) | 2750 (299-106000) |
|  | 3 | 33.4 (10.1-172) | 26.6 (8.85-148) | 8.78 (6.6-11.9) | 8.21 (5.78-11.9) | 6570 (525-190000) | 3930 (513-126000) |
|  | 4 | 38.5 (12-203) | 30 (9.91-168) | 9.2 (7.06-12.2) | 8.48 (6.17-13) | 9570 (787-287000) | 4660 (598-164000) |

Supplementary Table 2 shows the scores (RPS, DSS, SES) for the case forecasts at different time horizons for each country for both the baseline Endemic-Epidemic model without transmission between regions, covariates or seasonality and the full Endemic-Epidemic model. There is a clear deterioration in predictive performance with increasing forecast horizon for all countries for both models.

### Calibration: PIT histograms stratified by age groups

#### Czechia: age-stratified case forecasts

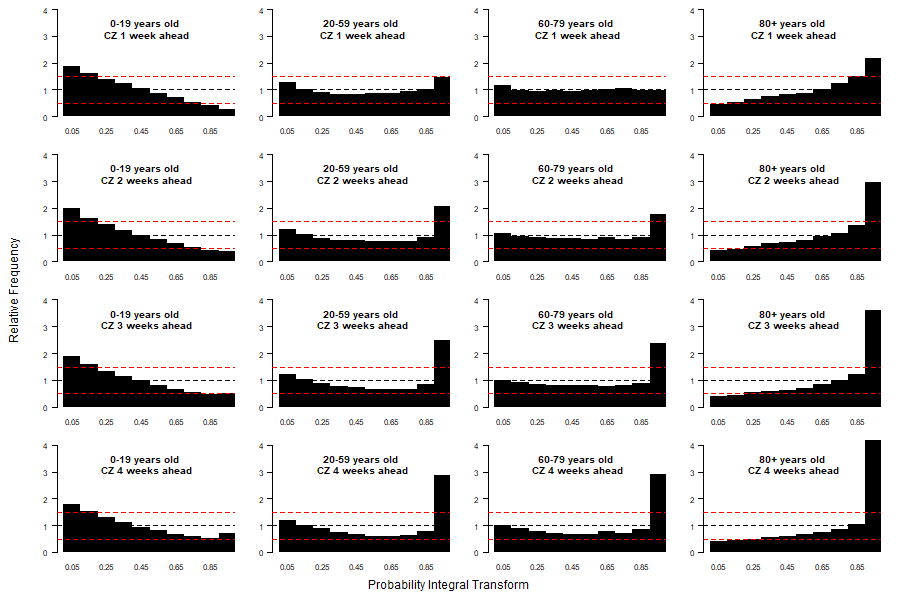
Supplementary Figure 2: PIT histograms showing the calibration of the daily case forecasts from the model for Czechia for 0-19 year-olds, 20-59 year-olds, 60-79 year-olds and 80+ year-olds (rows, top to bottom) for one-, two-, three- and four-week-ahead forecast horizons (columns, left to right) for 2nd May 2022 to 30th January 2023.

#### France: age-stratified case forecasts

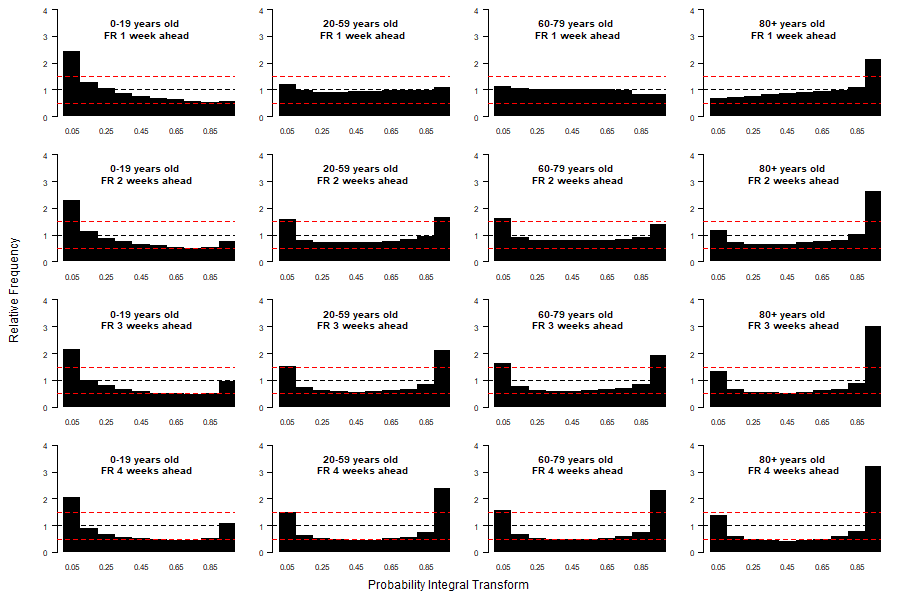

Supplementary Figure 3: PIT histograms showing the calibration of the daily case forecasts for France for 0-19 year-olds, 20-59 year-olds, 60-79 year-olds and 80+ year-olds (rows, top to bottom) for one-, two-, three- and four-week-ahead forecast horizons (columns, left to right) for 2nd May 2022 to 30th January 2023.

#### Czechia: age-stratified death forecasts

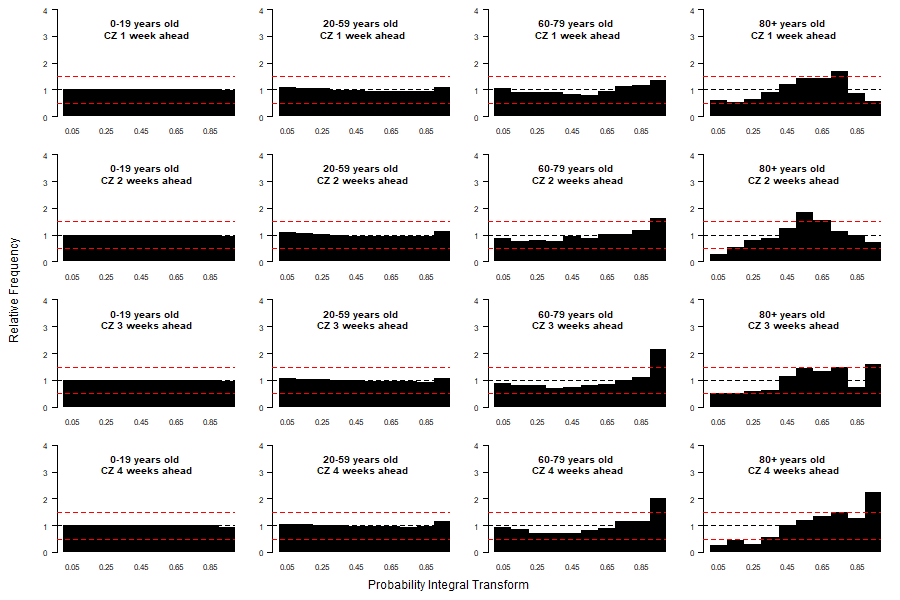
Supplementary Figure 4: PIT histograms showing the calibration of the weekly death forecasts from the model for Czechia for 0-19 year-olds, 20-59 year-olds, 60-79 year-olds and 80+ year-olds (rows, top to bottom) for one-, two-, three- and four-week-ahead forecast horizons (columns, left to right) for 2nd May 2022 to 30th January 2023.

#### France: age-stratified death forecasts

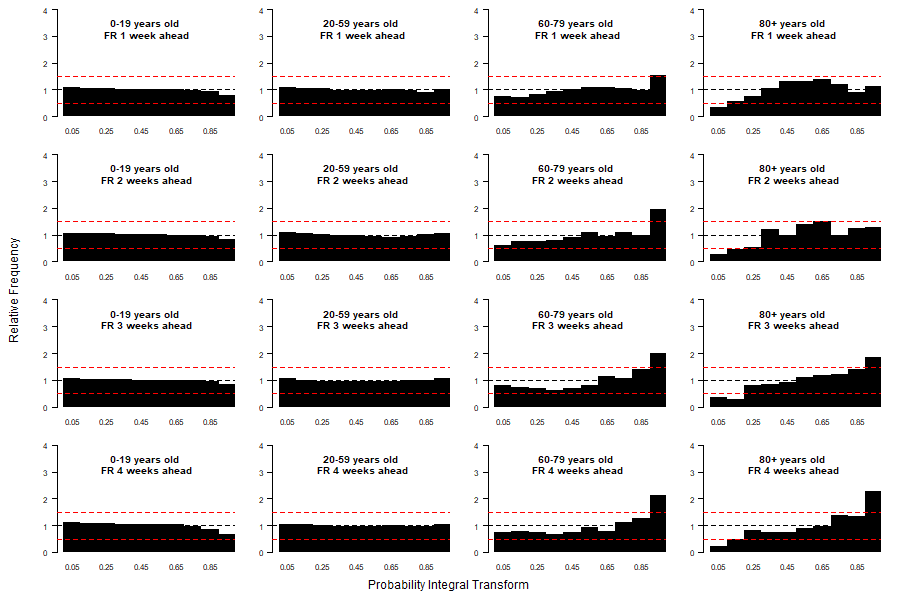

Supplementary Figure 5: PIT histograms showing the calibration of the weekly death forecasts from the model for France for 0-19 year-olds, 20-59 year-olds, 60-79 year-olds and 80+ year-olds (rows, top to bottom) for one-, two-, three- and four-week-ahead forecast horizons (columns, left to right) for 2nd May 2022 to 30th January 2023.

### Calibration of non-age stratified models

#### Country-level forecasts: case forecasts

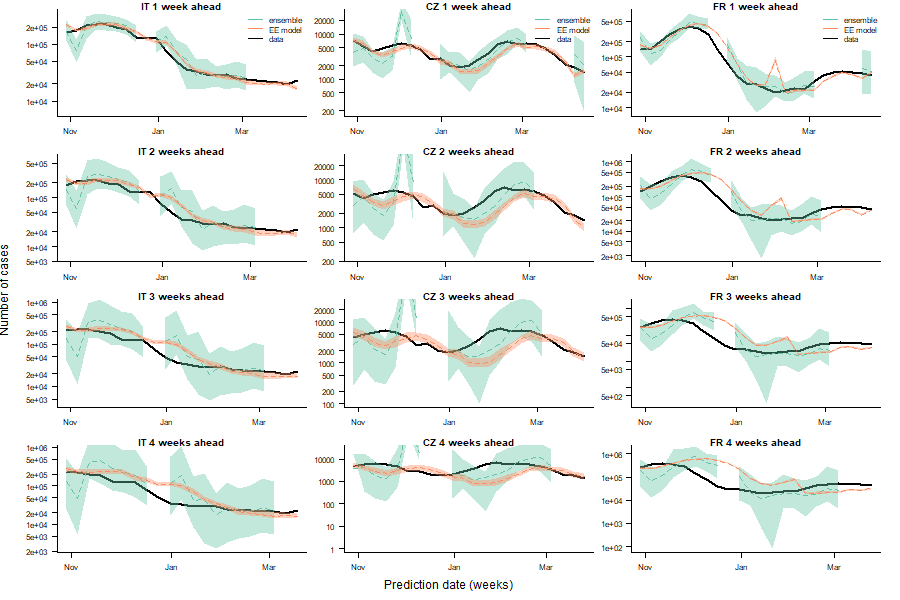

Supplementary Figure 6: Comparison of one-, two-, three- and four-week-ahead national-level case forecasts (rows, top to bottom) for Italy, Czechia and France (columns, left to right) from non-age-stratified version of our model (hhh4) with ensemble forecasts from the European COVID-19 Forecast Hub (ensemble) and observed cases (data) for 2nd May 2022 to 30th January 2023. Dashed lines show median forecasts, shaded regions the 95% prediction interval for the forecasts.

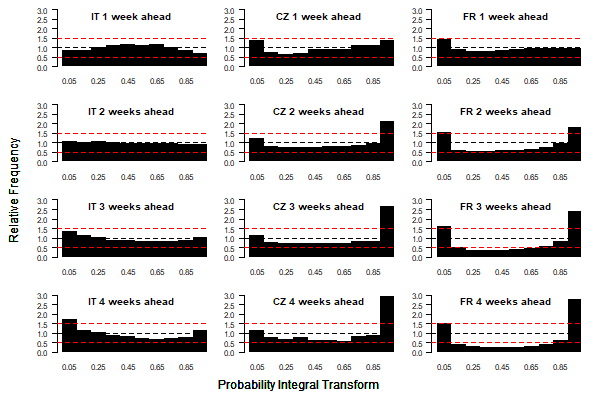

Supplementary Figure 7: PIT histograms showing the calibration of the daily case forecasts from the non-age-stratified version of the model for Italy, Czechia and France (columns, left to right) for one-, two-, three- and four-week-ahead forecast horizons (rows, top to bottom) for 2nd May 2022 to 30th January 2023.

#### Country-level forecasts: death forecasts

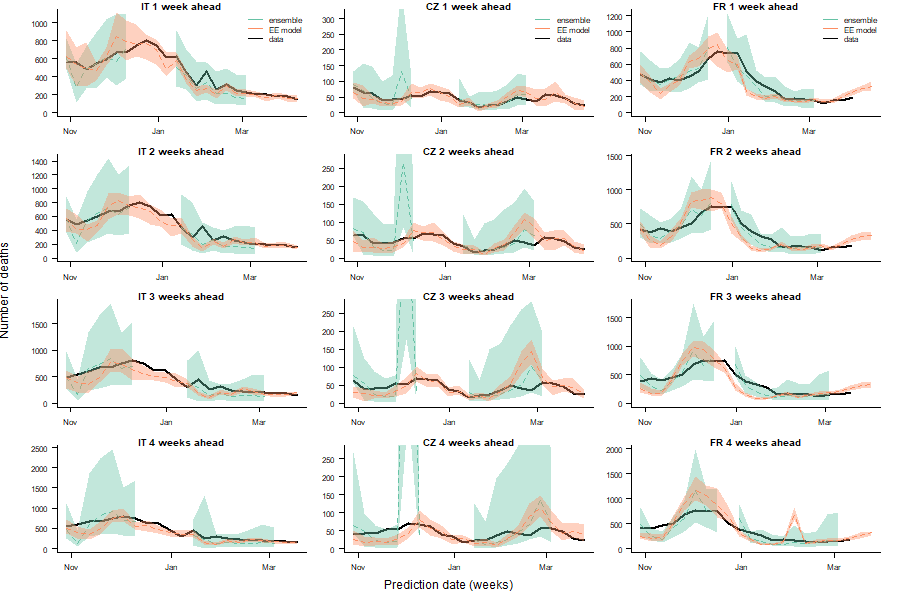

Supplementary Figure 8: Comparison of one-, two-, three- and four-week-ahead national-level death forecasts (rows, top to bottom) for Italy, Czechia and France (columns, left to right) from non-age-stratified version of our model (hhh4) with ensemble forecasts from the European COVID-19 Forecast Hub (ensemble) and observed cases (data) for 2nd May 2022 to 30th January 2023. Dashed lines show median forecasts, shaded regions the 95% prediction interval for the forecasts.

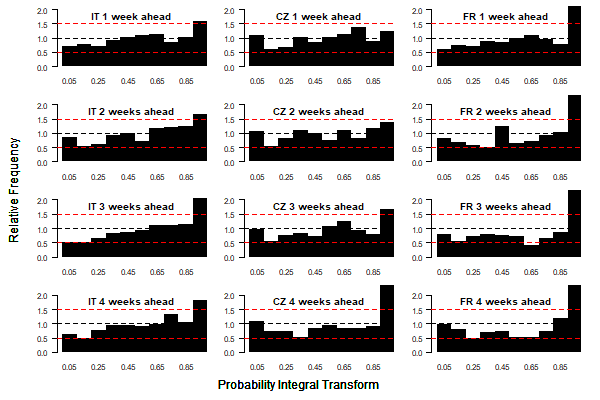

Supplementary Figure 9: PIT histograms showing the calibration of the weekly death forecasts from the non-age-stratified version of the model for Italy, Czechia and France (columns, left to right) for one-, two-, three- and four-week-ahead forecast horizons (rows, top to bottom) for 2nd May 2022 to 30th January 2023.

### References

1.	Béraud G, Kazmercziak S, Beutels P, Levy-Bruhl D, Lenne X, Mielcarek N, et al. The French Connection: The First Large Population-Based Contact Survey in France Relevant for the Spread of Infectious Diseases. Chuang JH, editor. PLoS One. 2015 Jul 15;10(7):e0133203.

2.	Prem K, Cook AR, Jit M. Projecting social contact matrices in 152 countries using contact surveys and demographic data. Halloran B, editor. PLoS Comput Biol. 2017 Sep 12;13(9):e1005697.

3.	[Eurostat. Methodology - Rural development [Internet]. [cited 2022 Jul 26]. Available from:](http://paperpile.com/b/XkGmkd/kW3I) <https://ec.europa.eu/eurostat/web/rural-development/methodology>

4.	[European Centre for Disease Prevention and Control. Data on SARS-CoV-2 variants in the EU/EEA [Internet]. [cited 2022 Jul 26]. Available from:](http://paperpile.com/b/XkGmkd/j9zH) <https://www.ecdc.europa.eu/en/publications-data/data-virus-variants-covid-19-eueea>

5.	Czado C, Gneiting T, Held L. Predictive model assessment for count data. Biometrics. 2009 Dec;65(4):1254–61.

6.	Bracher J, Ray EL, Gneiting T, Reich NG. Evaluating epidemic forecasts in an interval format. PLoS Comput Biol. 2021 Feb;17(2):e1008618.
